# SARS-CoV-2 Epidemiology on a Public University Campus in Washington State

**DOI:** 10.1101/2021.03.15.21253227

**Authors:** Ana A. Weil, Sarah L. Sohlberg, Jessica A. O’Hanlon, Amanda M. Casto, Anne W Emanuels, Natalie K. Lo, Emily P. Greismer, Ariana M. Magedson, Naomi C. Wilcox, Ashley E. Kim, Lewis Back, Christian D. Frazar, Ben Pelle, Thomas R. Sibley, Misja Ilcisin, Jover Lee, Erica L. Ryke, J. Chris Craft, Kristen M. Schwabe-Fry, Kairsten A. Fay, Shari Cho, Peter D. Han, Sarah J. Heidl, Brian A. Pfau, Melissa Truong, Weizhi Zhong, Sanjay R. Srivatsan, Katia F. Harb, Geoffrey S. Gottlieb, James P. Hughes, Deborah A. Nickerson, Christina M. Lockwood, Lea M. Starita, Trevor Bedford, Jay A. Shendure, Helen Y. Chu

**Affiliations:** Department of Medicine, Division of Allergy and Infectious Diseases, University of Washington, Seattle, WA USA; Department of Genome Sciences, University of Washington, Seattle, WA USA; Fred Hutchinson Cancer Research Center, Seattle, WA USA; Brotman Baty Institute for Precision Medicine, Seattle, WA USA; Department of Environmental Health and Safety, University of Washington, Seattle, WA USA; Department of Biostatistics, University of Washington, Seattle, WA USA

## Abstract

**Background:** Testing programs have been utilized as part of SARS-CoV-2 mitigation strategies on university campuses, and it is not known which strategies successfully identify cases and contain outbreaks.

**Objective:** Evaluation of a testing program to control SARS-CoV-2 transmission at a large university.

**Design:** Prospective longitudinal study using remote contactless enrollment, daily mobile symptom and exposure tracking, and self-swab sample collection. Individuals were tested if the participant was (1) exposed to a known case, developed new symptoms, or reported high-risk behavior, (2) a member of a group experiencing an outbreak, or (3) at baseline upon enrollment.

**Setting:** An urban, public university during Autumn quarter of 2020

**Participants:** Students, staff, and faculty.

**Measurements:** SARS-CoV-2 PCR testing was conducted, and viral genome sequencing was performed.

**Results:** We enrolled 16,476 individuals, performed 29,783 SARS-CoV-2 tests, and detected 236 infections. Greek community affiliation was the strongest risk factor for testing positive. 75.0% of positive cases reported at least one of the following: symptoms (60.8%), exposure (34.7%), or high-risk behaviors (21.5%). 88.1% of viral genomes (52/59) sequenced from Greek-affiliated students were genetically identical to at least one other genome detected, indicative of rapid SARS-CoV-2 spread within this group, compared to 37.9% (11/29) of genomes from non-Greek students and employees.

**Limitations:** Observational study.

**Conclusion:** In a setting of limited resources during a pandemic, we prioritized testing of individuals with symptoms and high-risk exposure during outbreaks. Rapid spread of SARS-CoV-2 occurred within outbreaks without evidence of further spread to the surrounding community. A testing program focused on high-risk populations may be effective as part of a comprehensive university-wide mitigation strategy to control the SARS-CoV-2 pandemic.

## INTRODUCTION

Universities are characterized by congregate living, in-person learning, and active social environments, all of which may contribute to rapid spread of infectious diseases. Between May and August, persons aged 20-29 accounted for over 20% of confirmed COVID-19 cases nationwide, and an even higher proportion in Washington state [1][2]. Numerous outbreaks on university campuses were observed early in the SARS-CoV-2 pandemic [3]. Surveillance strategies ranged from pooled testing of asymptomatic persons to wastewater analysis, with substantial heterogeneity across universities and no clear federal guidance [3–6]. Nationwide shortages of testing supplies and reagents have prevented many universities, including ours, from adopting a strategy of testing all members of the university community on a regular basis. Given these constraints, we sought to identify an approach that would permit early identification of cases and containment of spread [7][8].

## METHODS

### Setting

This study was conducted at a large public university in Seattle, Washington, composed of approximately 60,000 students and 30,000 faculty and staff [9][10]. In Autumn 2020, the university opened with hybrid in-person and remote classes, and residence halls were populated at a limited capacity [11]. The study population included students, staff, and faculty affiliated with the main campus and two smaller campuses located 15 and 35 miles from the main campus.

### Study Enrollment

Enrollment began September 24, 2020 during student resident move-in and continued during the study period. Eligibility criteria included living on-campus or within the main or satellite campus geographic area (approximately 100-mile radius), a valid university identification number, the ability to consent in English, and university attendance for class or work at least one day per month, either in-person or remotely. Exclusion criteria were living outside of the geographic area (i.e., people working remotely but living in another state) and age under 13 years. Participants completed informed consent and an online questionnaire that included baseline risk behaviors and demographic information. At enrollment, participants indicated a preference for either email or text communication. Study data were managed using Project REDCap software [12][13]. Individuals were stratified into risk tiers based on time spent on-campus, the number of individuals in their household, and university affiliation (Supplemental Table 1). Due to ongoing outbreaks in the Greek community, university and Greek community leadership strongly encouraged Greek student enrollment during the study.

### Attestations and Invitations to Test

Testing was offered for four reasons: (1) Attestation positivity; (2) Outbreaks; (3) Baseline surveillance; and (4) Holidays. Due to limited testing resources, we employed a hierarchical approach for invitations to test. "Attestation positive” study participants were prioritized, followed by outbreak invitations, and finally baseline and holiday invitations.

The daily attestation survey was used to determine testing eligibility. A participant was defined as “attestation positive” if they reported “yes” to the following questions: In the last 24 hours, have you 1) experienced new symptoms (“symptoms”), 2) been exposed to a COVID-19 positive individual (“exposure”), or 3) attended a high-risk gathering (“gathering”, defined as attending an indoor gathering of >10 people without social distancing or mask-wearing [Appendix 5]). Individuals who attested positive were offered testing, and repeat testing was offered up to every three days. If a “gathering” or “exposure” attestation was reported, a testing invite was delayed 48 hours to account for the incubation period. Outbreak testing was offered every three days to individuals of a community once an outbreak was identified by the university public health officials. Holiday testing was offered before and after the Thanksgiving break November 17th to December 6th.

### Testing Mechanisms

Testing was conducted at in-person kiosks or via mail-in swab kits (Appendix D). Participants affiliated with the main campus who received a testing invitation were offered in-person appointments of their choosing within 72 hours. Samples were collected by observed anterior nasal self-swab at on-campus kiosks. For participants affiliated with a satellite campuses or who indicated mobility restrictions precluding attendance at an on-campus kiosk, a self-testing kit was sent to and picked up from their residence using rapid courier services [14][15].

### Laboratory Methods and Results Reporting

Samples collected at kiosks were transported to the Northwest Genomics Center at the University of Washington and tested for SARS-CoV-2 using an RT-qPCR laboratory developed test (LDT). The laboratory is clinically certified to test for SARS-CoV-2 by the Washington State Department of Health Medical Testing Site. The RT-qPCR consists of assays for two SARS-CoV-2 targets in duplicate and the human marker RNase P across four multiplexed reactions (see Supplemental Text), [16]). A sample was considered positive if three or four replicates for RNase P had a cycle threshold (Ct) value <36 and SARS-CoV-2 had a cycle threshold (Ct) value <40. If only two SARS-CoV-2 replicate reactions were positive, the result was defined as inconclusive. Inconclusive results were regarded as low-positive results and participants were counselled identically to participants with a positive result [17]. If SARS-CoV-2 was not detected or detected in only one replicate, the test was considered negative. Samples were defined as failed and considered ‘never tested’ if RNase P was undetected in two or more reactions, or if there was a laboratory or operator error. Mid-study (November 18, 2020), we implemented an extraction-free testing method that yielded similar results, but Ct values from the two methods are not directly comparable [16].

Results were provided to participants through a research report hosted on a secure online portal that was accessed using a unique barcode identifier and date of birth. Cases were contacted by university, county, or state public health staff and contact tracing was initiated. Viral genome sequencing was attempted on all positive samples with a Ct value of 30 or less using a hybrid capture enrichment method [18] or a COVID-seq amplicon method (Illumina). Raw sequencing reads were processed using the Seattle Flu Study Assembly Pipeline (available on GitHub [19]). SARS-CoV-2 sequences were aligned and phylogenetic trees were constructed using the Nextstrain augur software [20]. Phylogenetic trees were visualized using the Nextstrain auspice software. All assembled genomes were publicly deposited to the Global Initiative on Sharing All Influenza Data (gisaid.org, [21]) and to Genbank immediately after data generation.

### University-Wide Prevention and Mitigation Strategy

Prior to and during Autumn quarter, the university deployed a communications campaign focused on masking, social distancing, handwashing, and disinfection of high-touch surfaces. Contact tracing was conducted by university public health officials for all students, staff, and faculty, except off-campus Greek community cases, which were handled by county public health. Isolation and quarantine housing was provided for infected students that lived in campus housing. University members also had access to free city SARS-CoV-2 testing sites or testing through their routine healthcare providers, outside of this research study.

### Statistical methods

Positive and inconclusive results were counted as cases. Confidence intervals were calculated using 95% confidence levels and p-values were considered statistically significant at alpha level of 0.05. Statistical testing for the comparison of averages was completed using Welch’s two sample t-tests. For multivariate regression, the reference group for race was White and non-Latinx/Hispanic for ethnicity.

COVID-19 cases were categorized as symptomatic, presymptomatic, asymptomatic, and possible asymptomatic. A case who tested positive/inconclusive was defined as “symptomatic” if they reported symptoms on their daily attestation survey within the seven days prior to testing or if they reported symptoms on the day of testing. “Presymptomatic” was defined as those who only reported symptoms in the week following testing, either on their daily attestation or follow-up survey. If no symptoms were reported before or after testing on the daily attestation or follow-up survey, they were considered “asymptomatic.” Participants who did not complete their daily attestations or the follow-up survey, and who reported no symptoms at the time of testing were labeled as “possible asymptomatic” cases.

A Generalized Estimating Equation with a logit link, robust variance, and an independent working correlation matrix was used to analyze risk factors for testing positive, allowing for dependence within individuals longitudinally. Odds ratios (OR) and their 95% confidence intervals were calculated adjusting for race, Latinx/Hispanic ethnicity, university or Greek affiliation, number of household members, attesting positive, mask wearing behavior, social distancing behavior, and on-campus frequency. All analyses were performed in R version 3.6.1

### Human Subjects

The University of Washington Institutional Review Board approved this study. All participants (or their guardians) provided written informed consent.

## RESULTS

Between September 24 and December 18, 2020, 16,476 individuals enrolled in the study, and 29,783 samples from 11,644 unique individuals were collected and tested for SARS-CoV-2 (see Supplemental Figure 1). Due to remote course instruction, many students were not living in the Seattle area. 25.5% (15,930/62,591) of matriculated students during Autumn quarter were enrolled in at least one course with in-person instruction, and 19.9% (8,204/41,296) of all matriculated undergraduates and 18.4% (2,719/14,765) of graduate students participated in the study [22–24]. More female (61.4% in the study vs. 54% in the student body), and White students (62.6% in the study vs. 40.8% in the student body; Supplemental Figure 2 and Supplemental Table 3) were enrolled. Of an estimated 4,100 Greek community students in the Seattle area at the time of the study, 2,672 (65.2%) were enrolled. The daily attestation survey was completed by a mean of 47.7% (6,203) participants per day (Figure 1A, 1B). Among participants attesting positive over the study period, 40.4% reported symptoms, 12.1% reported exposure, 36.4% reported gathering, and 11.1% reported multiple reasons. During the study, 409 (2.5%) participants opted to stop receiving daily attestation alerts, and were given the option of completing attestations on the study website. Results on preventative behaviors are listed in the Supplemental Results.

**Figure 1.**
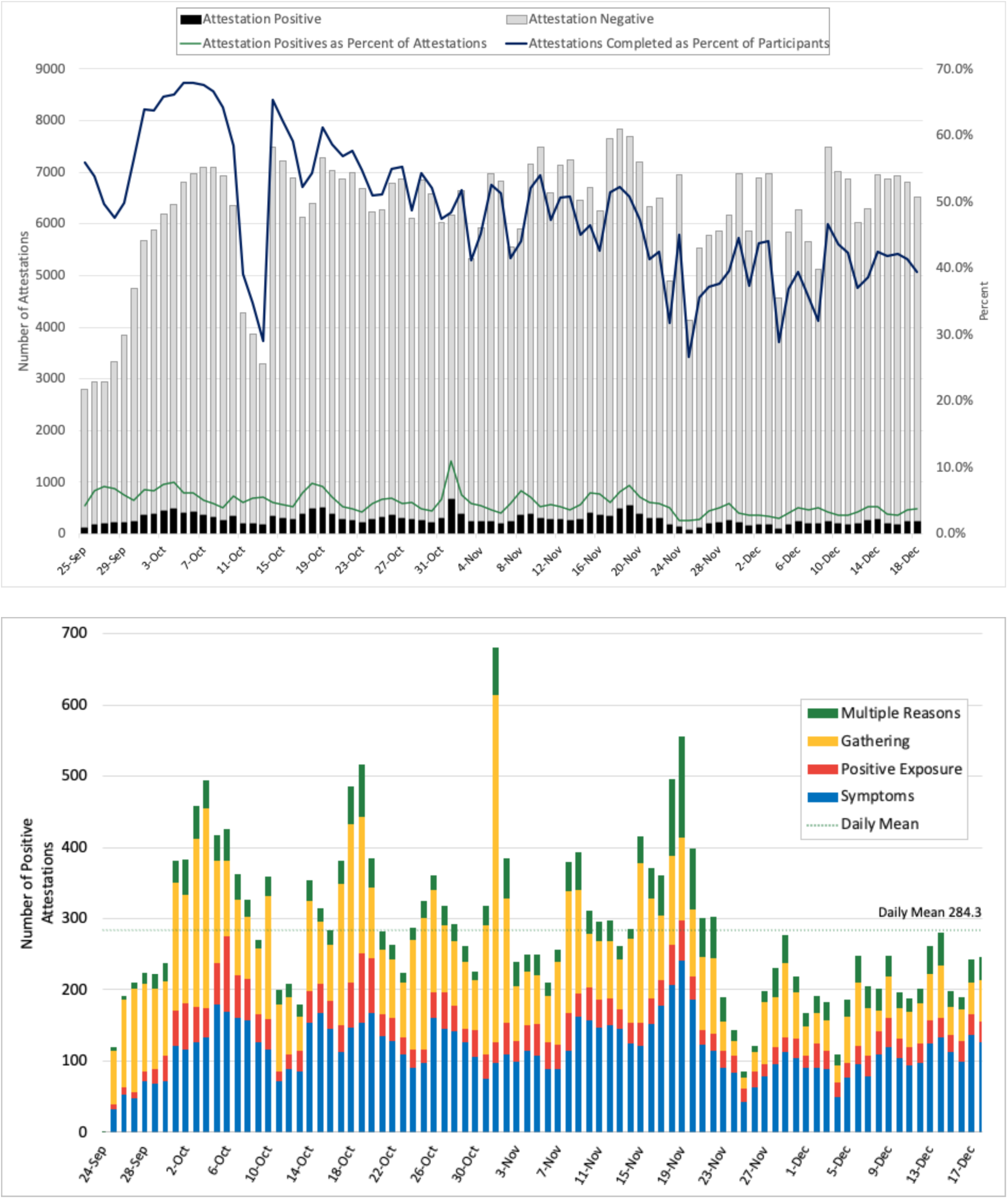
Daily attestation survey engagement over time. (A) Number of daily attestations completed daily during the study period. Between October 11-13, 2020 we experienced an outage of the text messaging service used to send daily attestation survey invites and this resulted in a reduced response rate. (B) Positive daily attestations stratified by reason for positive attestation. A marked increase in positive attestations was observed the day after Halloween, when 487 reported gathering, a 4.7-fold increase from the mean daily gathering attestation positive of 105.

### SARS-CoV-2 Testing Results

A total of 11,633 (70.6%) participants were tested at least once (Supplemental Table 2). Tests were resulted and available for participants to view online the day of sample collection (26.3%) or the following day (62.6%), and a minority of students received their results >24 hours after testing. 265 out of 29,783 samples (0.80%) tested positive or inconclusive for SARS-CoV-2 (Table 2). Among the 265 cases, there were 60.8% (61) symptomatic, 19.6% (52) presymptomatic, 3.4% (9) asymptomatic, and 16.2% (43) possible asymptomatic. Symptoms reported by SARS-CoV-2 result are reported in Supplemental Table 7. 92/256 (34.7%) of participants testing positive reported exposure to a known positive case. By group, the Greek community had 1.5% test positivity (1796/12045), on-campus dorm residents had 1.2% positivity (43/3507), and staff and faculty had 0.4% (19/4417) and 0.3% (4/1467) positivity, respectively. Test positivity by affiliation and race is shown in Supplemental Table 4. Overall, the university was aware of 745 positive SARS-CoV-2 individuals during Autumn quarter, of which 31.7% (236) were detected as part of this study [24]. For comparison of Ct values during Autumn quarter, we restricted comparisons to the time period before the change in testing methods, since methods used before and after November 18th were not comparable. Among samples collected from September 24th to November 18th, 2020, mean Ct values were higher in presymptomatic compared to symptomatic cases (28.7 vs. 24.2, p= 0.001) (Figure 2)

**Figure 2.**
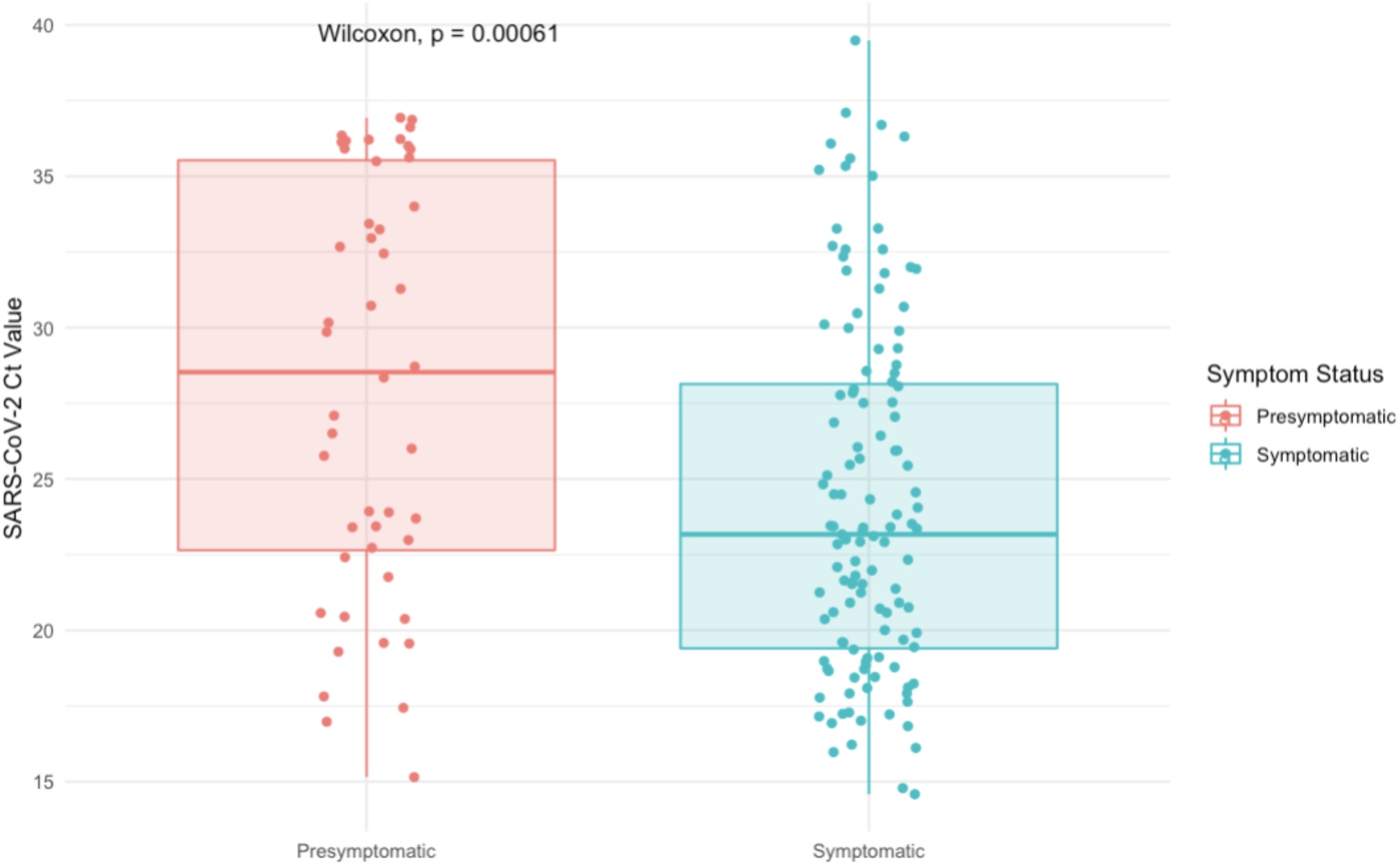
Comparison of Viral Load in Symptomatic (n=124) vs. Presymptomatic (n=48) Positive and Inconclusive Samples. Cycle threshold (Ct) for samples (each represented by one dot) tested using our protocol with nucleic acid extraction (before November 18) are shown here. Complete data is shown in Supplemental Figure 6. Box plots show the median values and 25^th^ and 75^th^ percentiles, with vertical lines demonstrating the range of values.

**Table 1.**
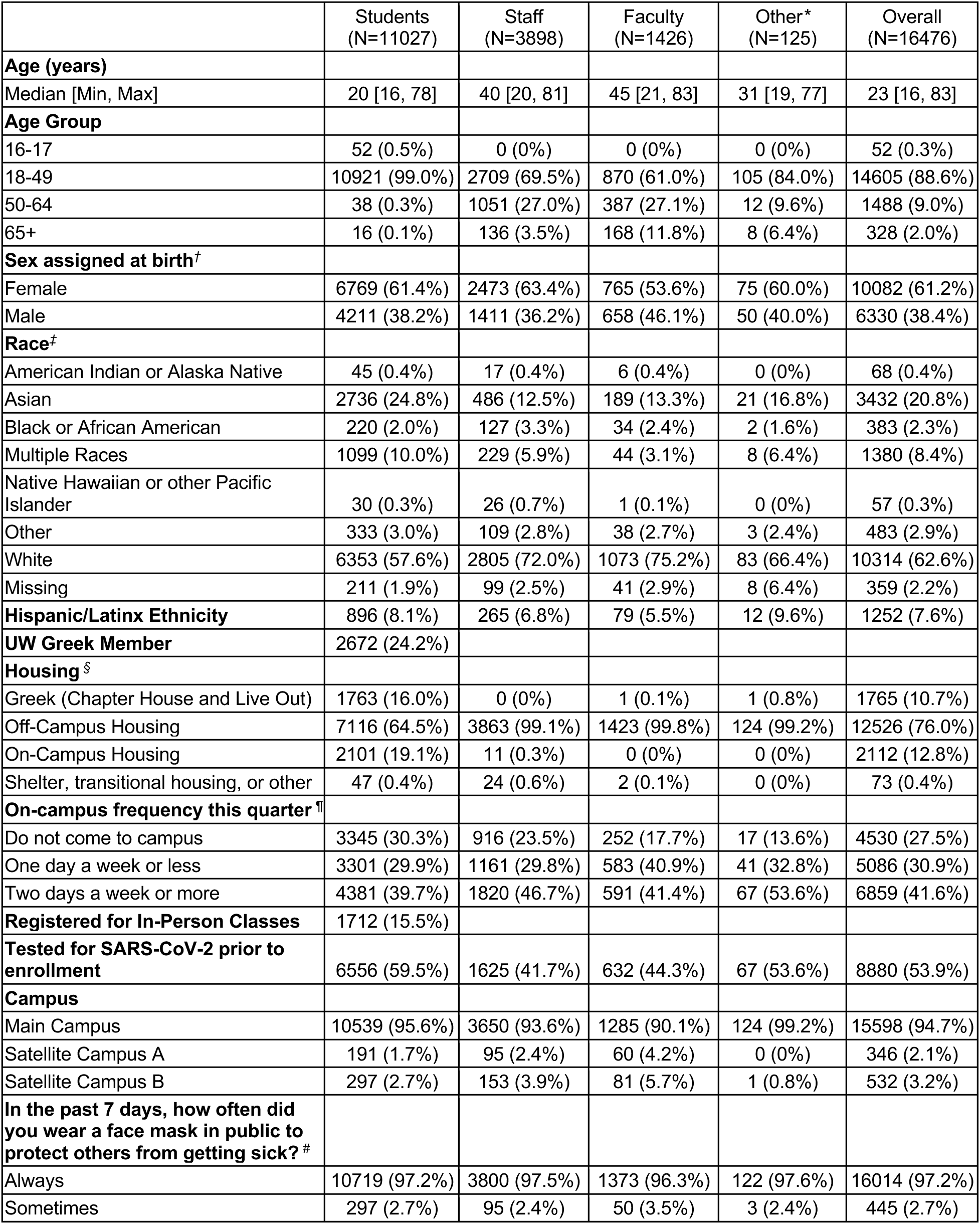
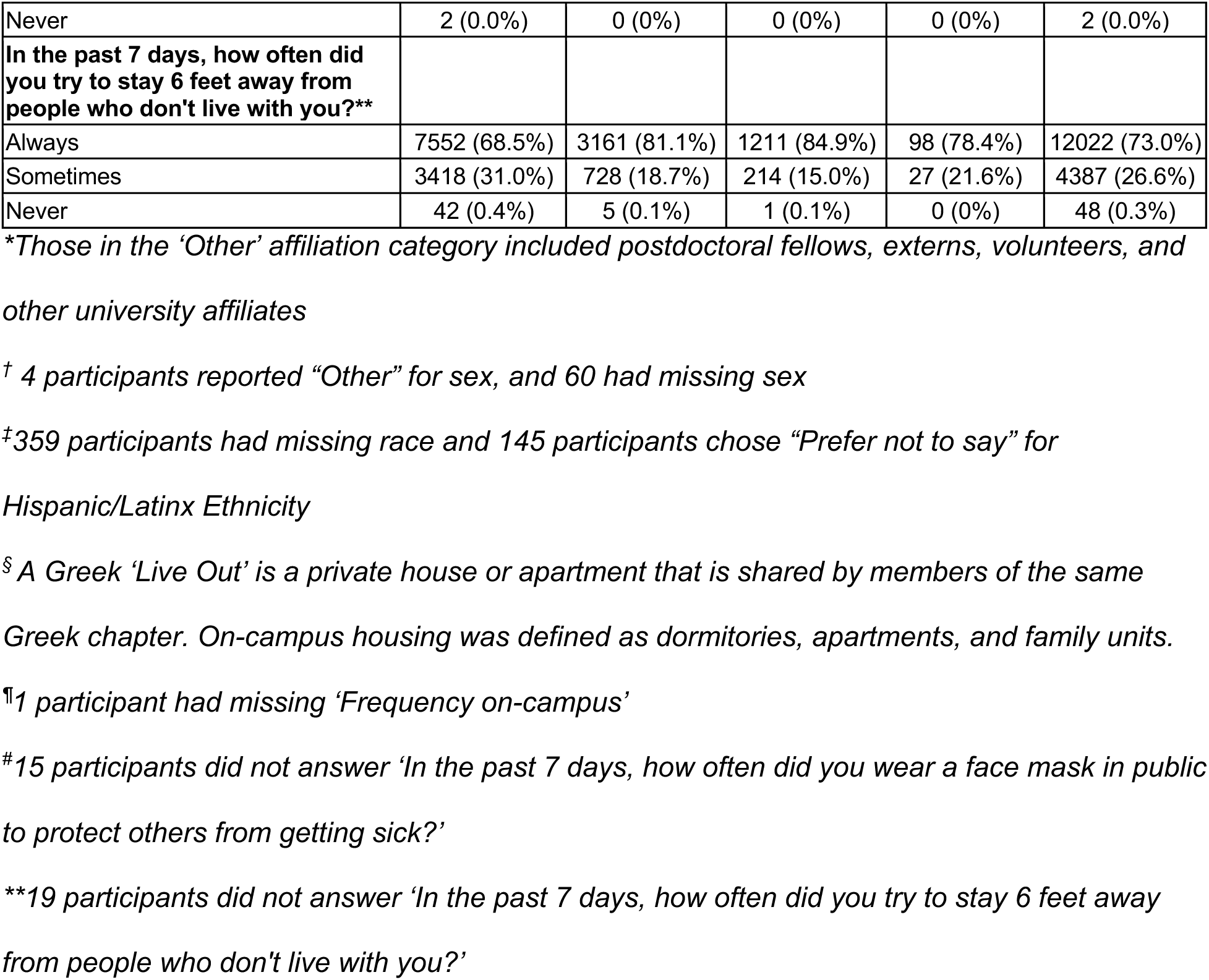
Sociodemographic Characteristics of Study Participants.

|  | Students<br>(N=11027) | Staff<br>(N=3898) | Faculty<br>(N=1426) | Other*<br>(N=125) | Overall<br>(N=16476) |
| --- | --- | --- | --- | --- | --- |
| **Age (years)** |  |  |  |  |  |
| Median [Min, Max] | 20 [16, 78] | 40 [20, 81] | 45 [21, 83] | 31 [19, 77] | 23 [16, 83] |
| **Age Group** |  |  |  |  |  |
| 16-17 | 52 (0.5%) | 0 (0%) | 0 (0%) | 0 (0%) | 52 (0.3%) |
| 18-49 | 10921 (99.0%) | 2709 (69.5%) | 870 (61.0%) | 105 (84.0%) | 14605 (88.6%) |
| 50-64 | 38 (0.3%) | 1051 (27.0%) | 387 (27.1%) | 12 (9.6%) | 1488 (9.0%) |
| 65+ | 16 (0.1%) | 136 (3.5%) | 168 (11.8%) | 8 (6.4%) | 328 (2.0%) |
| **Sex assigned at birth<sup>†</sup>** |  |  |  |  |  |
| Female | 6769 (61.4%) | 2473 (63.4%) | 765 (53.6%) | 75 (60.0%) | 10082 (61.2%) |
| Male | 4211 (38.2%) | 1411 (36.2%) | 658 (46.1%) | 50 (40.0%) | 6330 (38.4%) |
| **Race<sup>‡</sup>** |  |  |  |  |  |
| American Indian or Alaska Native | 45 (0.4%) | 17 (0.4%) | 6 (0.4%) | 0 (0%) | 68 (0.4%) |
| Asian | 2736 (24.8%) | 486 (12.5%) | 189 (13.3%) | 21 (16.8%) | 3432 (20.8%) |
| Black or African American | 220 (2.0%) | 127 (3.3%) | 34 (2.4%) | 2 (1.6%) | 383 (2.3%) |
| Multiple Races | 1099 (10.0%) | 229 (5.9%) | 44 (3.1%) | 8 (6.4%) | 1380 (8.4%) |
| Native Hawaiian or other Pacific Islander | 30 (0.3%) | 26 (0.7%) | 1 (0.1%) | 0 (0%) | 57 (0.3%) |
| Other | 333 (3.0%) | 109 (2.8%) | 38 (2.7%) | 3 (2.4%) | 483 (2.9%) |
| White | 6353 (57.6%) | 2805 (72.0%) | 1073 (75.2%) | 83 (66.4%) | 10314 (62.6%) |
| Missing | 211 (1.9%) | 99 (2.5%) | 41 (2.9%) | 8 (6.4%) | 359 (2.2%) |
| **Hispanic/Latinx Ethnicity** | 896 (8.1%) | 265 (6.8%) | 79 (5.5%) | 12 (9.6%) | 1252 (7.6%) |
| **UW Greek Member** | 2672 (24.2%) |  |  |  |  |
| **Housing<sup>§</sup>** |  |  |  |  |  |
| Greek (Chapter House and Live Out) | 1763 (16.0%) | 0 (0%) | 1 (0.1%) | 1 (0.8%) | 1765 (10.7%) |
| Off-Campus Housing | 7116 (64.5%) | 3863 (99.1%) | 1423 (99.8%) | 124 (99.2%) | 12526 (76.0%) |
| On-Campus Housing | 2101 (19.1%) | 11 (0.3%) | 0 (0%) | 0 (0%) | 2112 (12.8%) |
| Shelter, transitional housing, or other | 47 (0.4%) | 24 (0.6%) | 2 (0.1%) | 0 (0%) | 73 (0.4%) |
| **On-campus frequency this quarter<sup>¶</sup>** |  |  |  |  |  |
| Do not come to campus | 3345 (30.3%) | 916 (23.5%) | 252 (17.7%) | 17 (13.6%) | 4530 (27.5%) |
| One day a week or less | 3301 (29.9%) | 1161 (29.8%) | 583 (40.9%) | 41 (32.8%) | 5086 (30.9%) |
| Two days a week or more | 4381 (39.7%) | 1820 (46.7%) | 591 (41.4%) | 67 (53.6%) | 6859 (41.6%) |
| **Registered for In-Person Classes** | 1712 (15.5%) |  |  |  |  |
| **Tested for SARS-CoV-2 prior to enrollment** | 6556 (59.5%) | 1625 (41.7%) | 632 (44.3%) | 67 (53.6%) | 8880 (53.9%) |
| **Campus** |  |  |  |  |  |
| Main Campus | 10539 (95.6%) | 3650 (93.6%) | 1285 (90.1%) | 124 (99.2%) | 15598 (94.7%) |
| Satellite Campus A | 191 (1.7%) | 95 (2.4%) | 60 (4.2%) | 0 (0%) | 346 (2.1%) |
| Satellite Campus B | 297 (2.7%) | 153 (3.9%) | 81 (5.7%) | 1 (0.8%) | 532 (3.2%) |
| **In the past 7 days, how often did you wear a face mask in public to protect others from getting sick?<sup>#</sup>** |  |  |  |  |  |
| Always | 10719 (97.2%) | 3800 (97.5%) | 1373 (96.3%) | 122 (97.6%) | 16014 (97.2%) |
| Sometimes | 297 (2.7%) | 95 (2.4%) | 50 (3.5%) | 3 (2.4%) | 445 (2.7%) |
| Never | 2 (0.0%) | 0 (0%) | 0 (0%) | 0 (0%) | 2 (0.0%) |
| **In the past 7 days, how often did you try to stay 6 feet away from people who don't live with you?\*\*** |  |  |  |  |  |
| Always | 7552 (68.5%) | 3161 (81.1%) | 1211 (84.9%) | 98 (78.4%) | 12022 (73.0%) |
| Sometimes | 3418 (31.0%) | 728 (18.7%) | 214 (15.0%) | 27 (21.6%) | 4387 (26.6%) |
| Never | 42 (0.4%) | 5 (0.1%) | 1 (0.1%) | 0 (0%) | 48 (0.3%) |

*\*Those in the 'Other' affiliation category included postdoctoral fellows, externs, volunteers, and other university affiliates*
*† 4 participants reported "Other" for sex, and 60 had missing sex*
*‡359 participants had missing race and 145 participants chose "Prefer not to say" for Hispanic/Latinx Ethnicity*
*§ A Greek 'Live Out' is a private house or apartment that is shared by members of the same Greek chapter. On-campus housing was defined as dormitories, apartments, and family units.*
*¶1 participant had missing 'Frequency on-campus'*
*#15 participants did not answer 'In the past 7 days, how often did you wear a face mask in public to protect others from getting sick?'*
*\*\*19 participants did not answer 'In the past 7 days, how often did you try to stay 6 feet away from people who don't live with you?'*

**Table 2.** Characteristics of testing instances by SARS-CoV-2 result.

|  | Positive | Overall* | Percent Positivity (95% CI<sup>†</sup>) |
| --- | --- | --- | --- |
|  | (n=265) | (n=29783) |  |
| **Any symptoms** | 161 | 11116 | 1.4% (1.2 - 1.7) |
| **Attended large indoor gathering** | 57 | 6996 | 0.8% (0.6 - 1.1) |
| **Close contact with known positive** | 92 | 3877 | 2.4% (1.9 - 2.9) |
| **Baseline test** | 35 | 5700 | 0.6% (0.4 - 0.9) |
| **Outbreak test** | 83 | 5257 | 1.6% (1.3 - 2) |
| **Holiday test** | 3 | 1244 | 0.2% (0 - 0.7) |
| **Walk-in (no invite)** | 18 | 1788 | 1.0% (0.6 - 1.6) |
| **Risk tier** |  |  |  |
| Tier 1 | 220 | 19239 | 1.1% (1 - 1.3) |
| Tier 2 | 12 | 4291 | 0.3% (0.1 - 0.5) |
| Tier 3 | 33 | 6253 | 0.5% (0.4 - 0.7) |
| **Affiliation<sup>‡</sup>** |  |  |  |
| Student | 240 | 23802 | 1.0% (0.9 - 1.1) |
| Staff | 19 | 4417 | 0.4% (0.3 - 0.7) |
| Faculty | 4 | 1467 | 0.3% (0.1 - 0.7) |
| **Age group** |  |  |  |
| 13-17 | 2 | 153 | 1.3% (0.2 - 4.6) |
| 18-49 | 254 | 27783 | 0.9% (0.8 - 1) |
| 50-64 | 8 | 1548 | 0.5% (0.2 - 1) |
| 65+ | 1 | 299 | 0.3% (0 - 1.8) |
| **Sex assigned at birth** |  |  |  |
| Female | 180 | 19234 | 0.9% (0.8 - 1.1) |
| Male | 84 | 10474 | 0.8% (0.6 - 1) |
| **House members** |  |  |  |
| Lives alone | 16 | 2708 | 0.6% (0.3 - 1) |
| 2 people | 63 | 8051 | 0.8% (0.6 - 1) |
| 3-5 people | 34 | 7468 | 0.5% (0.3 - 0.6) |
| 6+ people | 152 | 11556 | 1.3% (1.1 - 1.5) |
| **UW Greek Member** | 176 | 12045 | 1.5% (1.3 - 1.7) |
| **Race** |  |  |  |
| American Indian or Alaska Native | 0 | 144 | 0.0% (0 - 2.5) |
| Asian | 32 | 4672 | 0.7% (0.5 - 1) |
| Black or African American | 2 | 460 | 0.4% (0.1 - 1.6) |
| Native Hawaiian or other Pacific Islander | 0 | 59 | 0.0% (0 - 6.1) |
| White | 191 | 20464 | 0.9% (0.8 - 1.1) |
| Multiple Races | 26 | 2808 | 0.9% (0.6 - 1.4) |
| Other | 10 | 707 | 1.4% (0.7 - 2.6) |
| **Latinx/Hispanic** | 35 | 2000 | 1.8% (1.2 - 2.4) |
| **Housing type** |  |  |  |
| Apartment | 32 | 7403 | 0.4% (0.3 - 0.6) |
| Dormitory/Residence Hall | 43 | 3507 | 1.2% (0.9 - 1.6) |
| House/condo/townhouse | 62 | 10433 | 0.6% (0.5 - 0.8) |
| Permanent supportive/ transitional housing | 0 | 6 | 0.0% (0 - 45.9) |
| Homeless Shelter | 0 | 5 | 0.0% (0 - 52.2) |
| UW Greek Chapter House | 104 | 6145 | 1.7% (1.4 - 2) |
| UW Greek Live Out | 24 | 2168 | 1.1% (0.7 - 1.6) |
| Other | 0 | 116 | 0.0% (0 - 3.1) |
| **Overall** | 265 | 29783 | 0.9% (0.8 - 1) |

*\*Overall\* includes 60 samples that were never tested*
*†Confidence intervals generated using Clopper-Pearson exact methods*
*‡86 'Other' and 15 'Volunteer' affiliation*

### Risk Factors for SARS-CoV-2 Infection

On multivariate analysis, Greek affiliation had the strongest association with test positivity (OR 2.71, 95% CI: 1.84-4.00, p <0.001). Latinx/Hispanic ethnicity (OR 2.12, 95% CI:1.28-2.18, p=0.002) and positive attestations (OR 1.86, 95% CI:1.43-2.41, p<0.001) were also risk factors for positivity (Supplemental Table 6). Reported frequency of hand washing, mask-wearing, and social distancing were not associated with positivity.

### Greek Community Outbreak

Thirty cases were identified in the Greek community during the first 10 days of the study, which prompted outbreak testing. Test positivity trends in the Greek community demonstrated a unique epidemiologic curve compared to the non-Greek students, employees, and the county (Figure 3A, [25]). Outbreaks within Greek houses were concurrent, but with unique individual timelines, and involved both fraternities and sororities (Figure 3B). 68.3% of Greek members reported sharing a living space with 6 or more people, compared to only 14.0% of non-Greek students. During 37 days of outbreak testing, serial testing frequently identified individuals who tested negative several times prior to testing positive (Figure 3C).

**Figure 3.**
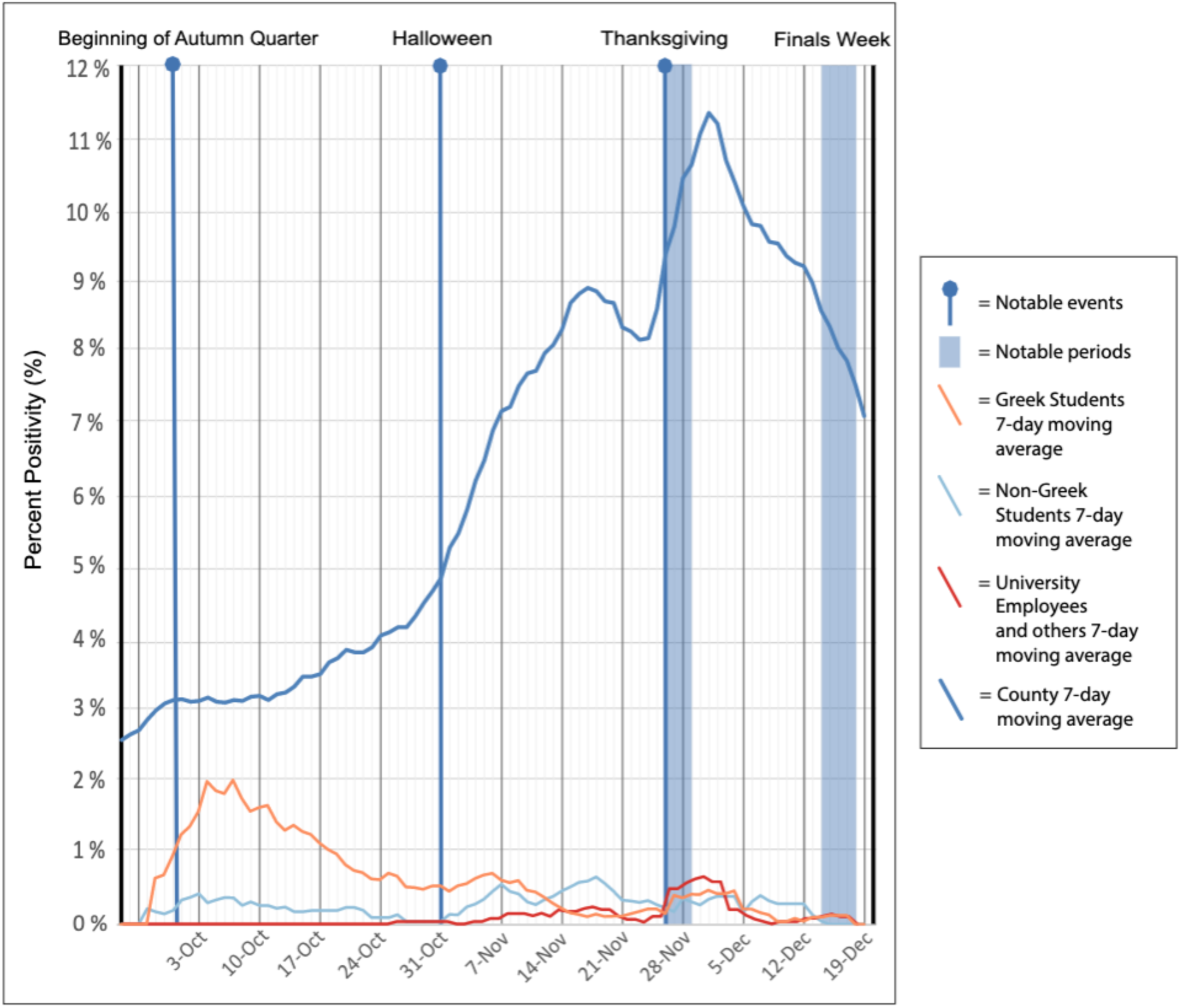
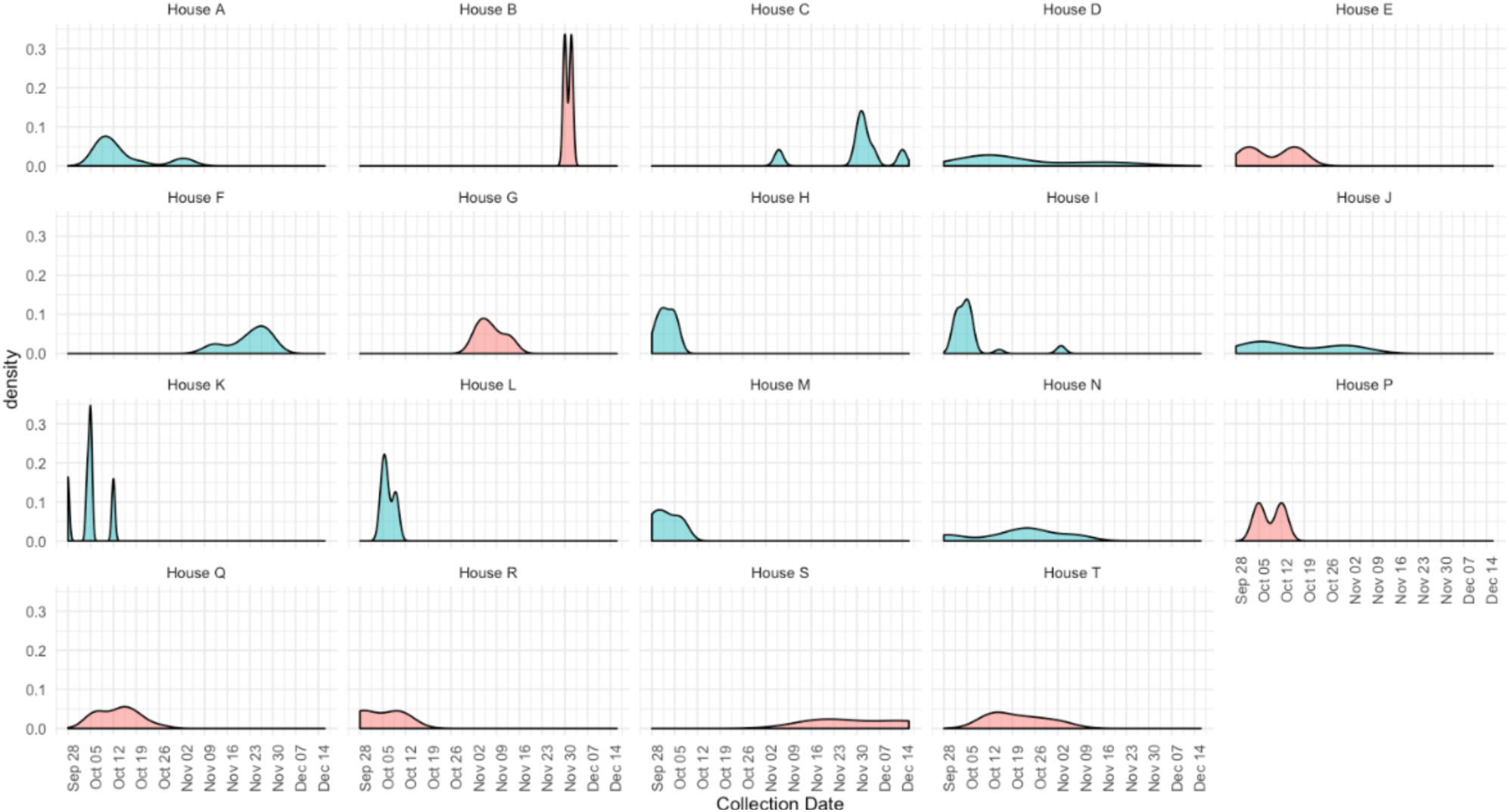
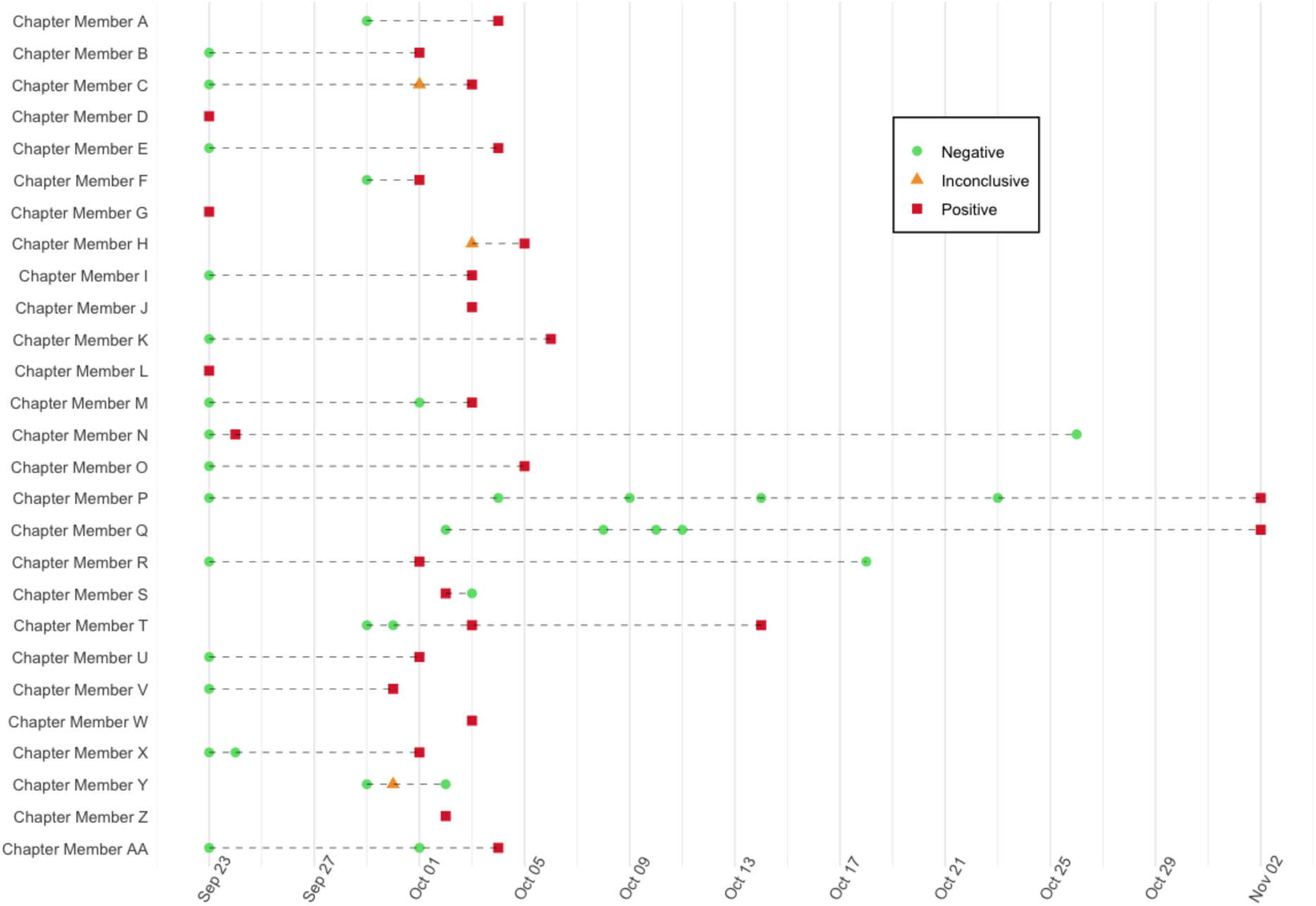
Dynamics of a Greek community outbreak. (A) Percent positivity over time of university groups and the surrounding county. The employee category includes university staff and faculty, and the Greek-affiliated students category includes all Greek-affiliated students including those living in Greek houses, Greek off-campus housing, and Greek-affiliated dorm residents. (B) Greek Chapter-level SARS-CoV-2 outbreak dynamics. Counts of cases identified by chapter during Autumn quarter. Sororities are shown in blue and the fraternities in red. Chapters with no infections detected (n=20) or <=2 infections detected (n=5) are not shown. (C) Example of chapter-level individual SARS-CoV-2 outbreak dynamics within one Greek chapter (ending on November 2nd). Lines represent individual study participants tested multiple times, and dots signify a single test.

### SARS-CoV-2 Molecular Epidemiology

High coverage genomes were generated for 88 SARS-CoV-2 samples collected from unique individuals between September 27th and November 28th. 59 were collected from Greek- affiliated students, 24 from non-Greek-affiliated students, and 5 from faculty or staff. In Figure 4A, a phylogenetic tree of 1700 SARS-CoV-2 genomes collected statewide, including the 88 samples from this study, is shown. Multiple study samples appear in each of the four major clades (20A, 20B, 20C, 20G) circulating in the county and state during this timeframe.

**Figure 4:**
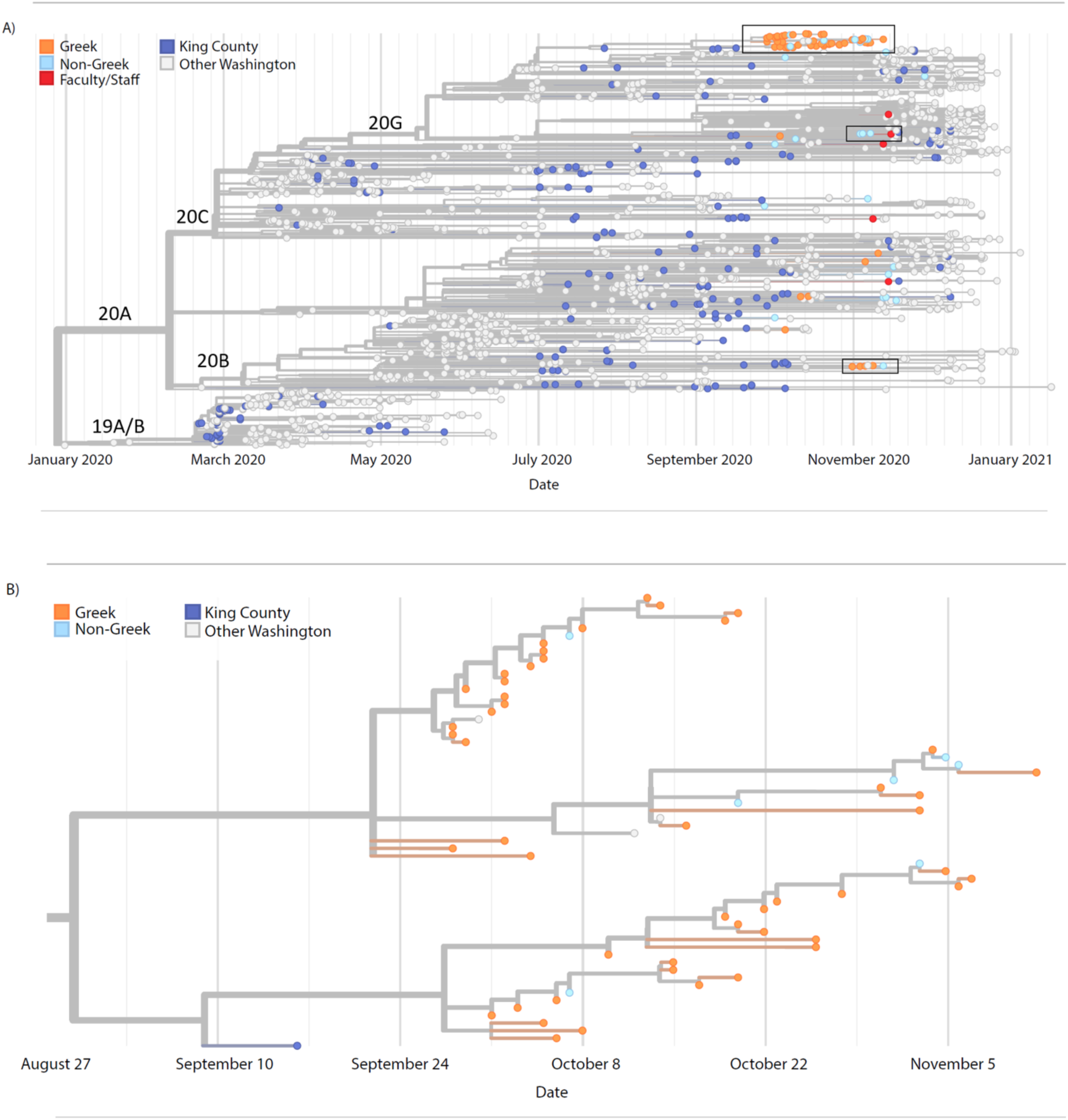
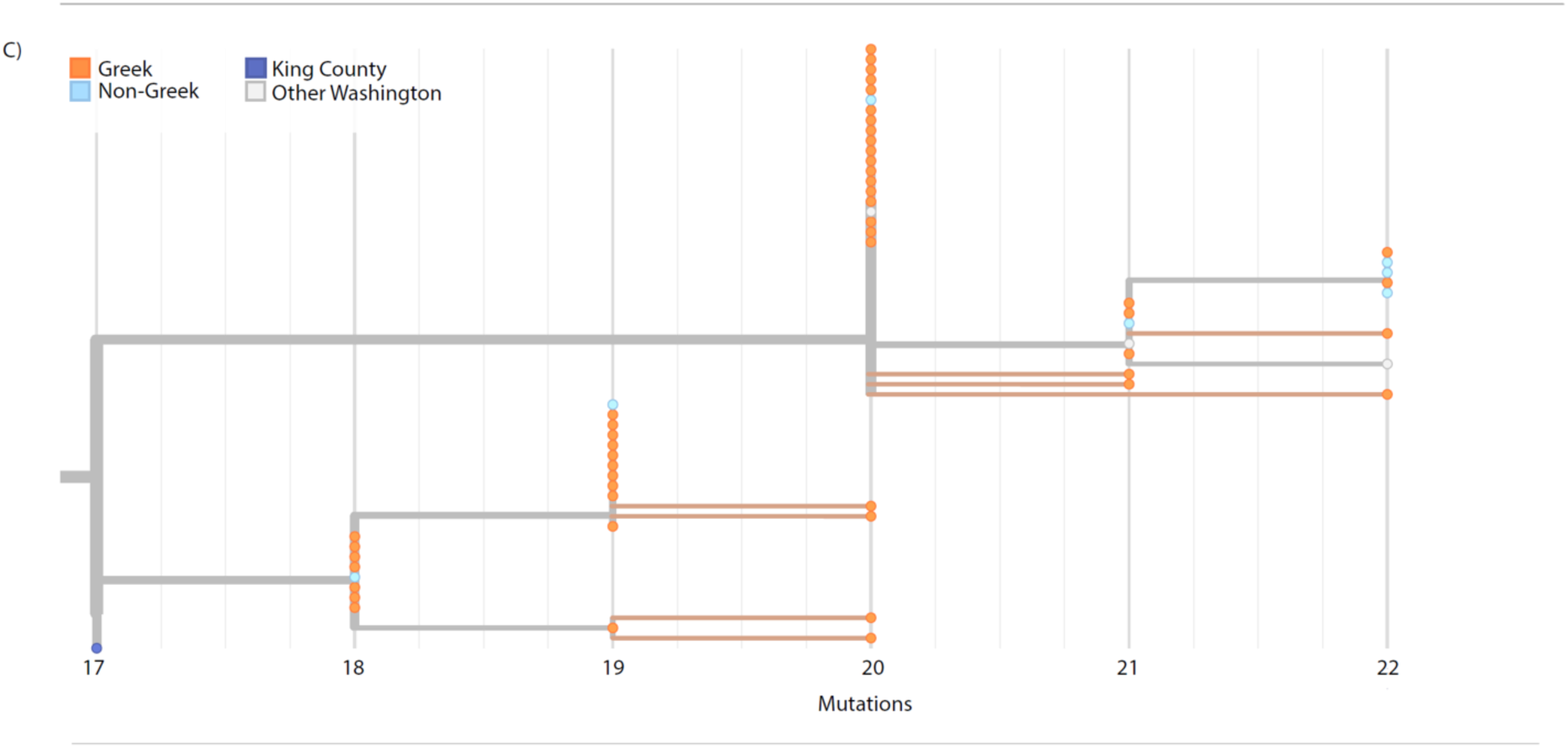
Molecular Epidemiology of a university outbreak. A) Phylogenetic tree of SARS- CoV-2 samples from Washington, including 88 samples from this study. Included here are all SARS-CoV-2 genomes from Washington collected on or after September 25, 2020, a random subsample of 1000 Washington samples collected prior to September 25, and the Wuhan/Hu-1 reference genome. Samples are positioned on the X axis by date of collection. B) Detail of a cluster of university genomes organized horizontally by collection date, and C) by divergence, or the number of genetic changes relative to SARS-CoV-2 reference genome.

Most viral genomes from this study (56/88, 63.6%) grouped into one large cluster that included genomes from 49 Greek-affiliated and 7 non-Greek-affiliated students (Clade 20G, large black box Figures 4A, B, C, and Supplemental Figure 3). This cluster also included genomes from four samples collected elsewhere in the state, but outside of our study. Samples in this cluster were collected between September 27th and November 12th, and all samples in this cluster collected prior to October 7th originated from Greek-affiliated students.

Closer inspection of this cluster (Figure 4B, C) demonstrates two sub-clusters (branch support values are 0.94 for larger sub-cluster and 0.78 for smaller sub-cluster). Samples within clusters are closely related with a maximum pairwise distance between any two samples in the same sub-cluster of four single nucleotide changes. Molecular clock estimates place the common ancestor of the larger cluster at September 22nd (95% CI: September 1^st^-29th) and the smaller cluster at September 27th (95% CI: September 20th - October 4th). These two dates occur just prior to the sharp increase in cases observed in our study among Greek students, which peaked on approximately October 7th-8th (Figure 3A). The last sample mapping to either of these sub- clusters was collected November 12th. This date roughly coincides with the end of the Greek outbreak as measured by the percent positivity rate (Figure 3A). Among the over 500 viral genomes collected state-wide after November 12th, including 9 collected by this study, none have descended from the viruses responsible for the Greek community olutbreak.

Two smaller clusters of viral genomes from this study are shown in Figure 4A (two small boxes) and Supplemental Figures 4, 5. One cluster contains five study genomes, four from Greek-affiliated students and one from a non-Greek student. This cluster has a most recent common ancestor dating to October 31st (95% CI: October 17^th^-November 1st). The second contains three genomes from non-Greek-affiliated students and one faculty/staff member with a most recent common ancestor dating to November 3rd (95% CI: October 11th-November 16th). Viral genomes from Greek-affiliated students were most likely to cluster with other study samples; 88.1% of genomes from Greek-affiliated students were genetically identical to at least one other study sample while only 45.8% of samples from non-Greek students and 0% of samples from faculty/staff were identical to another sample.

## DISCUSSION

Our large-scale COVID-19 longitudinal cohort study of students, faculty, and staff at a university campus allocated testing based on estimated risk of infection, rather than mass surveillance, a strategy that could be utilized in a pandemic setting with resource limitations. Most cases were identified through daily attestation surveys, and participants reporting a recent exposure to a case had the highest positivity rate of 2.4%, followed by 1.4% for participants reporting symptoms. We found that baseline testing had a much lower positivity rate of 0.6%, and identified only 15% of cases. Consistent with our findings, random testing of asymptomatic people at the University of Pittsburgh yielded a positivity rate of only 0.4% [5]. In contrast, at Duke University, surveillance testing of asymptomatic persons identified half of all cases [4]. Several college campus outbreaks have been linked to spread in communities and high-risk groups such as long-term care facility residents, and campus outbreaks are known indicators of broader community transmission [27][28]. In our study, SARS-CoV-2 test positivity rates in the university community were substantially lower than in the surrounding areas. This is likely due in part to increased testing availability, particularly for those without symptoms. However, it is also possible that focused testing of high-risk groups prevented and mitigated campus-wide outbreaks.

Outbreak testing was an essential part of our testing strategy. In our study, Greek affiliation was the most important risk factor for testing positive, and more than two-thirds of the cases detected in this study were in Greek-affiliated students. A study conducted at an Arkansas university, in which 91% of identified SARS-CoV-2 cases occurred in Greek-affiliated students, previously implicated this group as high-risk for contracting COVID-19 [29]. We observed cases in more than half of our university’s Greek chapters, and the genomic analysis and outbreak dynamics indicate that transmission occurred at a rapid pace both within and among fraternities and sororities. However, later in the quarter, the decreased test positivity rate among this group became comparable to the rates observed among the non-Greek participants. We believe this decline was driven in part by our aggressive testing strategy in Greek-affiliated students, which identified pre- and asymptomatic cases, and was followed by effective contact tracing. Genomic analysis of a subset of the SARS-CoV-2 samples collected in this study revealed major differences between those from Greek community members and those from non-Greek community members. In particular, sequenced samples from Greek students were much more likely than samples from non-Greek students or from faculty/staff to form clusters. Our phylogenetic analysis suggests that most cases of SARS-CoV-2 detected in Greek-affiliated students were the result of campus-related transmission while the genetic diversity observed in non-Greek students or faculty/staff cases were more likely to have been acquired in the community. While most sequenced samples from Greek students formed a single large cluster that may have resulted from a single introduction event of the virus into the Greek community, the sub-division of this large cluster into at least two distinct sub-clusters suggests that viral spread among Greek students occurred as a series of separate transmission events. The hypothesis that social behaviors, rather than housing arrangements, drove the Greek outbreak is supported by our analyses that rapid spread occurred not only within, but also among Greek houses.

We did not observe evidence of Greek-related virus spread into the wider community. Given that a limited number of SARS-CoV-2 genomes were sequenced, this does not rule out “spillover” of virus from the Greek outbreak into the outside community, but to date, none of the sequenced SARS-CoV-2 samples collected state-wide appear to descend from the Greek outbreak virus.

In addition to members of the Greek community, participants who identified as Latinx or Hispanic experienced increased rates of test positivity. This is consistent with COVID-19 incidence rates in Washington State, where Hispanic individuals represented 13% of state residents, but 33% of COVID-19 cases [26]. This highlights the need for more targeted and equitable distribution of testing and contact tracing resources in this high-risk population.

Testing and behavioral interventions are cost-effective interventions to control outbreaks on college campuses; however, prioritization of testing is critical, and scarcity of laboratory supplies were present prior to and during our study [30]. We employed several mechanisms to conserve testing resources. First, we used the daily attestation surveys to screen for symptoms predictive of SARS-CoV-2 infection [31] to prioritize tests for those at highest risk. Second, we used mass produced swabs that are shipped dry and eluted in PCR-friendly buffer to allow an extraction-free RT-qPCR method. By avoiding the need for limiting reagents such as transport media and RNA extraction kits, we avoided supply chain challenges, reduced the price per test, and increased the speed of our testing pipeline, enabling us to maintain a 24-hour turnaround time on average while increasing scale.

Limitations of this study include that our online format of consent, enrollment, and daily attestation surveys increased participant engagement but was a barrier to participation for individuals with limited technological literacy or access. Additionally, study materials were available in English only. While on-campus testing through this program was often the most convenient testing option, the data included here is not comprehensive in describing on-campus cases, since other testing mechanisms were also available. The race and ethnicity representation in the study population was likely skewed by a university campaign to target, enroll, and test members of the Greek community. Our ability to speculate on the patterns related to Greek community transmission dynamics are impacted both by the availability of other testing mechanisms for members of the university community and inability to sequence all study samples.

We report here a strategy for SARS-CoV-2 testing on a large university campus using contactless, rapid enrollment and self-administered testing during Autumn quarter 2020. Most infections were detected in the Greek community, and this group experienced distinct genomic and epidemiologic dynamics compared to other university communities and the surrounding area. This evidence suggests that testing those engaged in high-risk activities, in combination with testing people experiencing symptoms, may be sufficient to stop on-campus transmission and prevent community spread in a setting of limited resources during a pandemic.

## Data Availability

Data and code used for analyses may be available upon request.

## ACKNOWLEDGEMENTS

We would like to thank the study participants. We also thank UW Environmental Health & Safety COVID-19 Prevention & Response team (Sheryl Schwartz, Megan Gourley, David Coomes, Casey Adams), UW administration and Incident Command leadership group (Margaret Shepherd, Derek Fulwiler, David Hotz, Josh Gana, Erik Johnson, Pamela Schreiber), Chu Lab and BBI HCT team (Jody Sicuro, Dylan McDonald, Devon McDonald, Madison Contor, Cooper Marshall, Lincoln Pothan, Taylor Wilson, Zack Acker, and Evan Sarantinos), Dr. Janet Englund, Dr. Timothy Uyeki, Dr. Michael Jackson, and Dr. Roy Burstein. We gratefully acknowledge the authors and the originating and submitting laboratories of the sequences from GISAID’s EpiFlu Database, on which this research is based.

## SUPPLEMENTARY MATERIAL

**Supplemental Figure 1.**
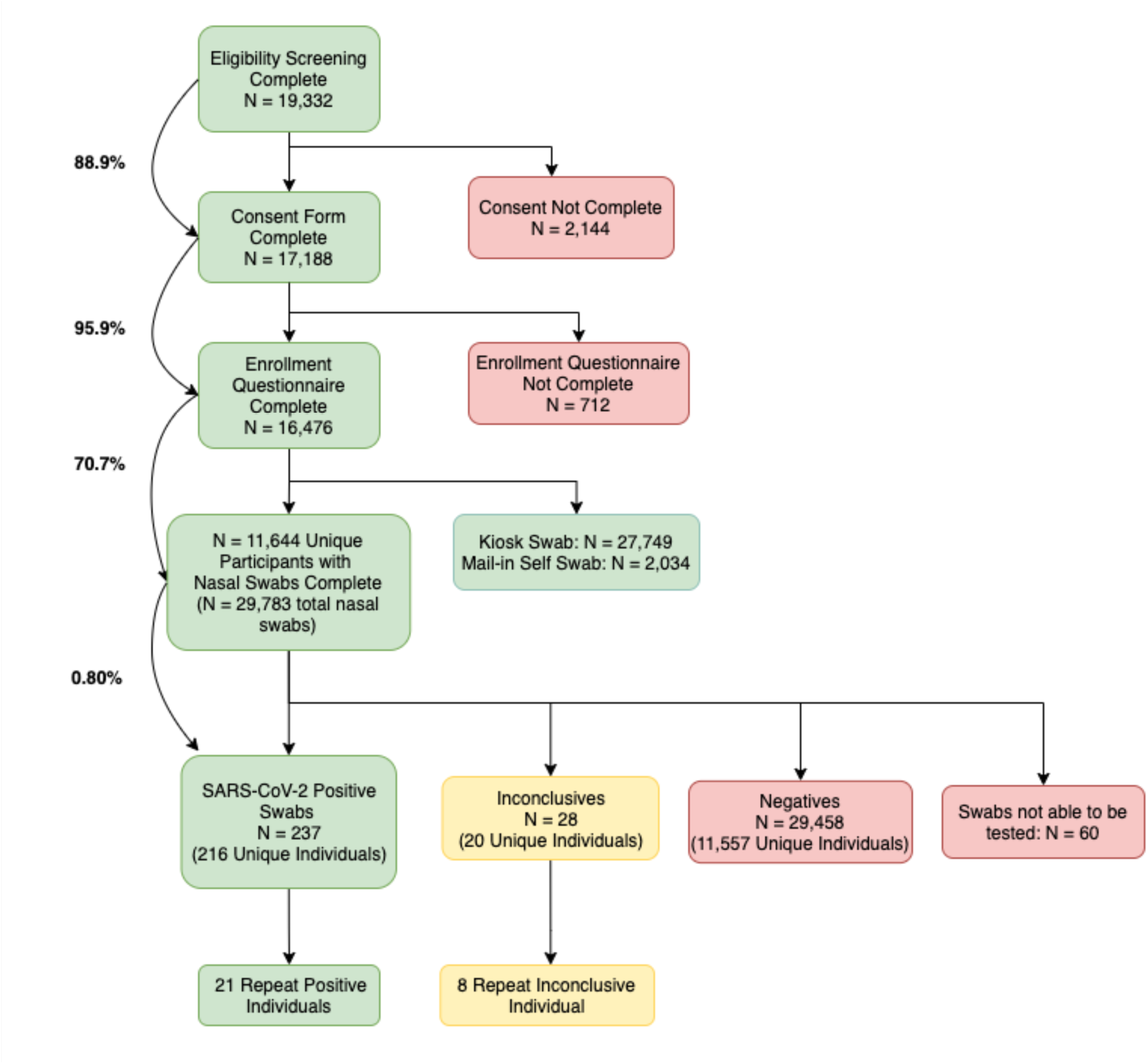
Participant study flow. Flow of study procedures from the participant’s view, beginning with the eligibility screening and ending with the test results. 11.1% of people who started the eligibility screening process did not complete the consent form, but nearly all (95.9%) of those who completed the consent form continued to complete the enrollment questionnaire.

**Supplemental Table 1.** **Tier Criteria for study participants.**

| Tier Level | Criteria |
| --- | --- |
| Tier 1 | Students living in on-campus housing, including dormitories/residence halls |
|  | Students affiliated with the Greek community or living in Greek housing |
|  | Students living in a household of 4 or more members |
| Tier 2 | Students on-campus two or more days per week that live in a household of 3 members or less |
|  | Faculty, Staff, and Volunteers of the main Campus that are on-campus two or more days per week |
| Tier 3 | All other enrollees |

**Supplemental Table 2.** **Enrollment and Testing Status by Tier**

|  | **Tier 1**<br>**(n=6638)** | **Tier 2**<br>**(n=3313)** | **Tier 3**<br>**(n=6525)** | **Overall**<br>**(n=16476)** |
| --- | --- | --- | --- | --- |
| **Testing Status** |  |  |  |  |
| Never Tested | 1326 (20.0%) | 758 (22.9%) | 2759 (42.3%) | 4843 (29.4%) |
| Tested at least once | 5312 (80.0%) | 2555 (77.1%) | 3766 (57.7%) | 11633<br>(70.6%) |
| **Number of swabs per**<br>**person** |  |  |  |  |
| Mean (SD) | 2.89 (3.14) | 1.29 (1.31) | 0.954 (1.27) | 1.80 (2.40) |
| Median [Min, Max] | 2.00 [0.00,<br>21.0] | 1.00 [0.00,<br>11.0] | 1.00 [0.00,<br>11.0] | 1.00 [0.00,<br>21.0] |

**Supplemental Figure 2.**
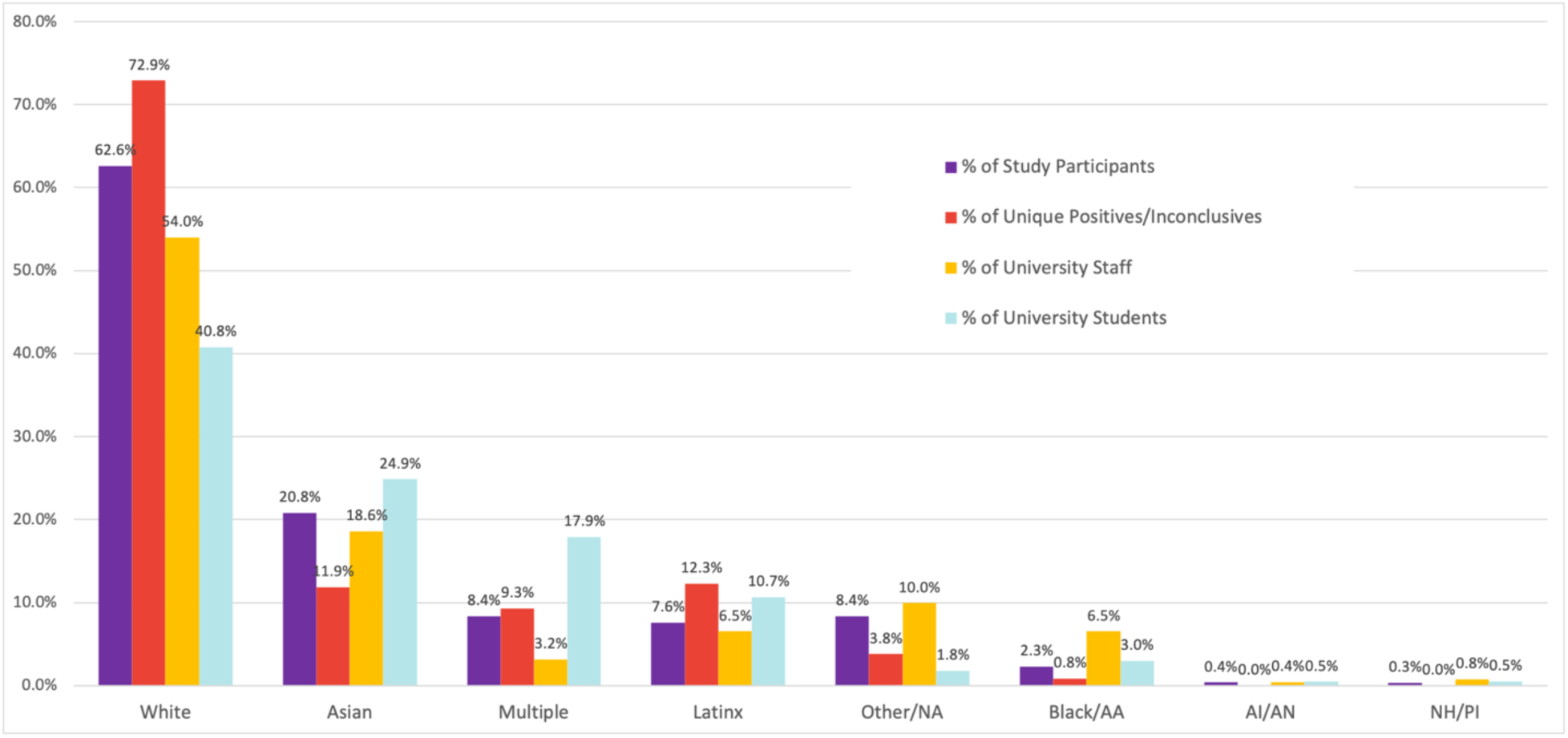
Enrollment and testing representation by race and ethnicity. The blue bars in Supplemental Figure 2 represent the percentage of study enrollees by self-identified race and ethnicity (Hispanic/Latinx) category. The purple bars represent the percentage of the study’s enrollees, red bars represent the percentage of the study’s unique individuals who tested positive or inconclusive, while the yellow and turquoise bars represent the percentage of UW overall student body and staff, respectively. Data on race and ethnicity categories of faculty were not available. Multiple = selected multiple race categories. AA = African American. AI/AN = American Indian or Alaska Native. NH/PI = Native Hawaiian or Pacific Islander.

**Supplemental Table 3.** **Race/Ethnicity in Study Population**

|  | Greek Student<br>(N=2672) | Non-Greek<br>Student<br>(N=8356) | Staff/Faculty<br>(N=5448) | Overall*<br>(N=16476) |
| --- | --- | --- | --- | --- |
| **Race** |  |  |  |  |
| American Indian or<br>Alaska Native | 10 (0.4%) | 35 (0.4%) | 23 (0.4%) | 68 (0.4%) |
| Asian | 324 (12.1%) | 2413 (28.9%) | 695 (12.8%) | 3432 (20.8%) |
| Black or African<br>American | 33 (1.2%) | 187 (2.2%) | 163 (3.0%) | 383 (2.3%) |
| Multiple Races | 286 (10.7%) | 813 (9.7%) | 281 (5.2%) | 1380 (8.4%) |
| Native Hawaiian or other<br>Pacific Islander | 8 (0.3%) | 22 (0.3%) | 27 (0.5%) | 57 (0.3%) |
| Other | 41 (1.5%) | 292 (3.5%) | 150 (2.8%) | 483 (2.9%) |
| White | 1944 (72.8%) | 4409 (52.8%) | 3961 (72.7%) | 10314 (62.6%) |
| **Latinx/Hispanic** | 167 (6.2%) | 729 (8.7%) | 356 (6.5%) | 1252 (7.6%) |

*\*359 missing race and 145 missing Hispanic/Latinx ethnicity*

**Supplemental Table 4.** **Test Positivity by University Affiliation and Race/Ethnicity**

|  |  | SARS-CoV-2 Test Result |  |  |  |
| --- | --- | --- | --- | --- | --- |
|  | Race Category | Positive | Inconclusive | Negative | Total |
| Student | American Indian or Alaska Native | 0 (0%) | 0 (0%) | 115 (100%) | 115 |
|  | Asian | 25 (0.6%) | 1 (0.0%) | 4009 (99.2%) | 4040 |
|  | Black or African American | 2 (0.7%) | 0 (0%) | 274 (98.6%) | 278 |
|  | Multiple Races | 21 (0.8%) | 3 (0.1%) | 2451 (98.8%) | 2481 |
|  | Native Hawaiian or other Pacific Islander | 0 (0%) | 0 (0%) | 37 (100%) | 37 |
|  | Other | 4 (0.8%) | 1 (0.2%) | 516 (98.5%) | 524 |
|  | White | 159 (1.0%) | 20 (0.1%) | 15833 (98.8%) | 16031 |
| Staff | American Indian or Alaska Native | 0 (0%) | 0 (0%) | 23 (100%) | 23 |
|  | Asian | 5 (1.1%) | 0 (0%) | 434 (98.6%) | 440 |
|  | Black or African American | 0 (0%) | 0 (0%) | 139 (98.6%) | 141 |
|  | Multiple Races | 1 (0.4%) | 0 (0%) | 277 (99.6%) | 278 |
|  | Native Hawaiian or other Pacific Islander | 0 (0%) | 0 (0%) | 22 (100%) | 22 |
|  | Other | 4 (2.8%) | 0 (0%) | 139 (97.2%) | 143 |
|  | White | 8 (0.2%) | 1 (0.0%) | 3237 (99.3%) | 3260 |
| Faculty | American Indian or Alaska Native | 0 (0%) | 0 (0%) | 6 (100%) | 6 |
|  | Asian | 0 (0%) | 1 (0.6%) | 180 (99.4%) | 181 |
|  | Black or African American | 0 (0%) | 0 (0%) | 40 (97.6%) | 41 |
|  | Multiple Races | 1 (2.4%) | 0 (0%) | 41 (97.6%) | 42 |
|  | Other | 1 (2.6%) | 0 (0%) | 37 (97.4%) | 38 |
|  | White | 1 (0.1%) | 0 (0%) | 1099 (99.5%) | 1105 |
| Other | Asian | 0 (0%) | 0 (0%) | 11 (100%) | 11 |
|  | Multiple Races | 0 (0%) | 0 (0%) | 7 (100%) | 7 |
|  | Other | 0 (0%) | 0 (0%) | 2 (100%) | 2 |
|  | White | 2 (2.9%) | 0 (0%) | 66 (97.1%) | 68 |

\* 469 participants who had race missing were excluded from the table. 'Overall' includes 60 samples that were never tested

**Supplemental Table 5.**
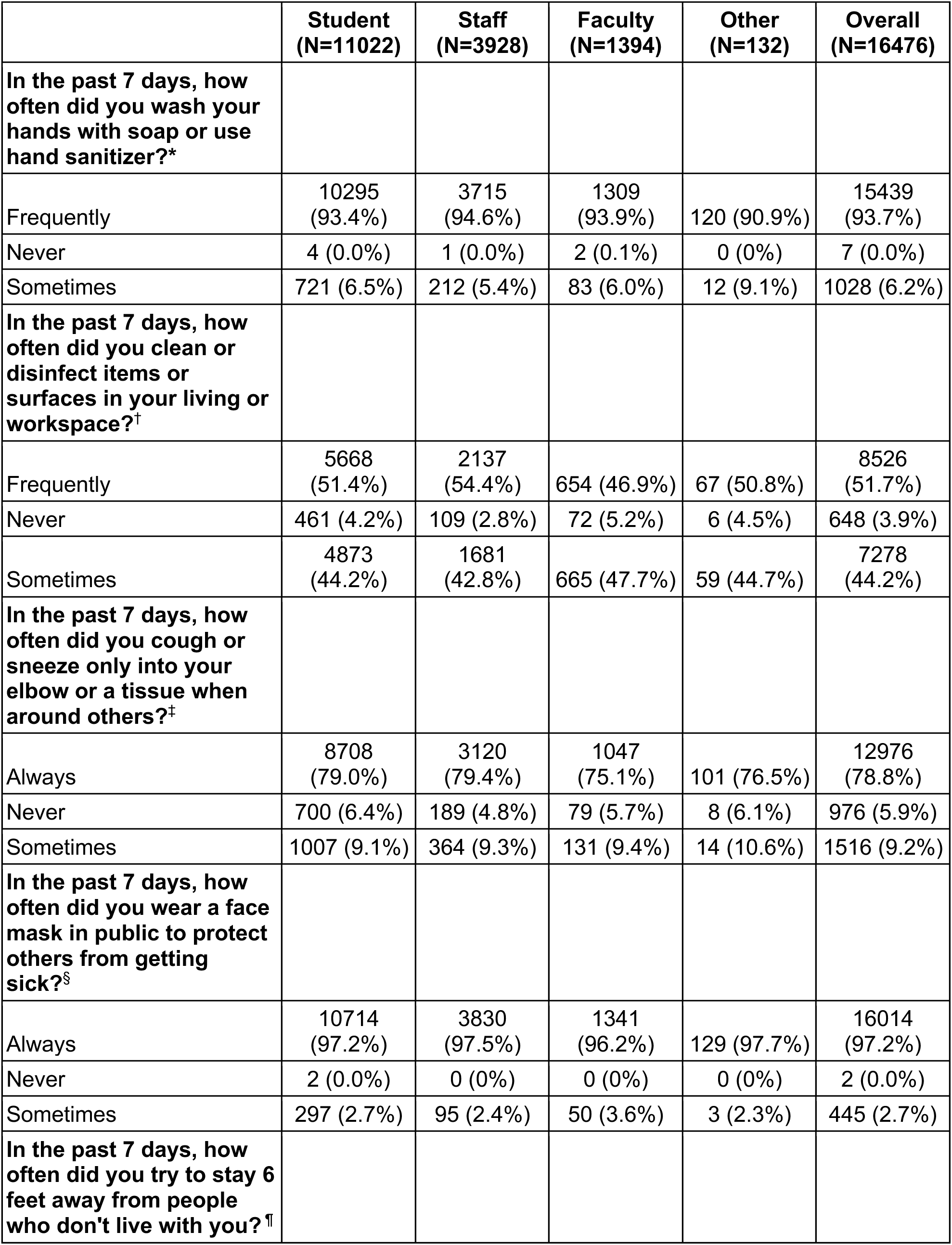
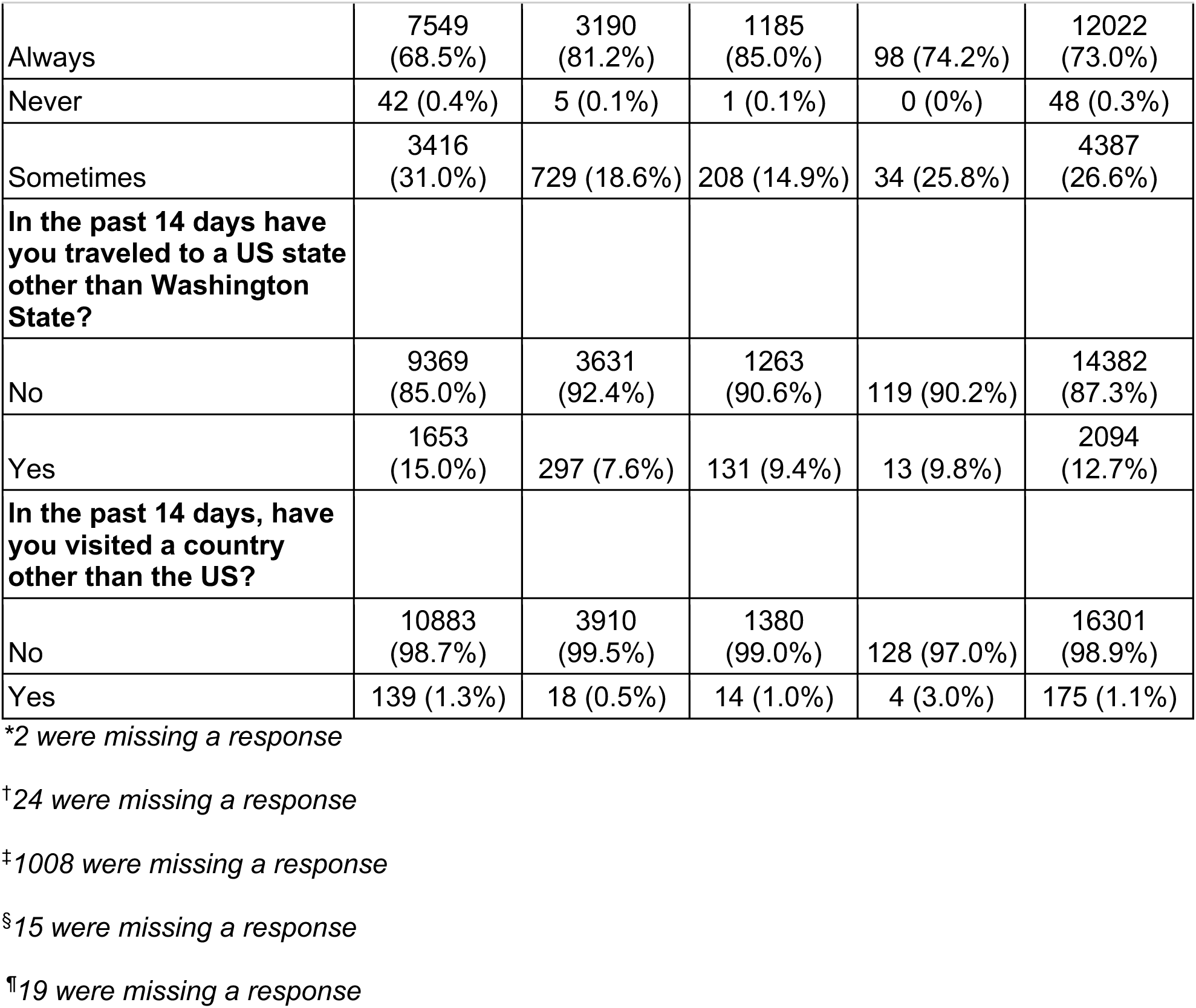
**Behavior and Travel Questions at Time of Enrollment**

|  | **Student**<br>**(N=11022)** | **Staff**<br>**(N=3928)** | **Faculty**<br>**(N=1394)** | **Other**<br>**(N=132)** | **Overall**<br>**(N=16476)** |
| --- | --- | --- | --- | --- | --- |
| **In the past 7 days, how often did you wash your hands with soap or use hand sanitizer?\*** |  |  |  |  |  |
| Frequently | 10295<br>(93.4%) | 3715<br>(94.6%) | 1309<br>(93.9%) | 120 (90.9%) | 15439<br>(93.7%) |
| Never | 4 (0.0%) | 1 (0.0%) | 2 (0.1%) | 0 (0%) | 7 (0.0%) |
| Sometimes | 721 (6.5%) | 212 (5.4%) | 83 (6.0%) | 12 (9.1%) | 1028 (6.2%) |
| **In the past 7 days, how often did you clean or disinfect items or surfaces in your living or workspace?†** |  |  |  |  |  |
| Frequently | 5668<br>(51.4%) | 2137<br>(54.4%) | 654 (46.9%) | 67 (50.8%) | 8526<br>(51.7%) |
| Never | 461 (4.2%) | 109 (2.8%) | 72 (5.2%) | 6 (4.5%) | 648 (3.9%) |
| Sometimes | 4873<br>(44.2%) | 1681<br>(42.8%) | 665 (47.7%) | 59 (44.7%) | 7278<br>(44.2%) |
| **In the past 7 days, how often did you cough or sneeze only into your elbow or a tissue when around others?‡** |  |  |  |  |  |
| Always | 8708<br>(79.0%) | 3120<br>(79.4%) | 1047<br>(75.1%) | 101 (76.5%) | 12976<br>(78.8%) |
| Never | 700 (6.4%) | 189 (4.8%) | 79 (5.7%) | 8 (6.1%) | 976 (5.9%) |
| Sometimes | 1007 (9.1%) | 364 (9.3%) | 131 (9.4%) | 14 (10.6%) | 1516 (9.2%) |
| **In the past 7 days, how often did you wear a face mask in public to protect others from getting sick?§** |  |  |  |  |  |
| Always | 10714<br>(97.2%) | 3830<br>(97.5%) | 1341<br>(96.2%) | 129 (97.7%) | 16014<br>(97.2%) |
| Never | 2 (0.0%) | 0 (0%) | 0 (0%) | 0 (0%) | 2 (0.0%) |
| Sometimes | 297 (2.7%) | 95 (2.4%) | 50 (3.6%) | 3 (2.3%) | 445 (2.7%) |
| **In the past 7 days, how often did you try to stay 6 feet away from people who don't live with you?¶** |  |  |  |  |  |
| Always | 7549<br>(68.5%) | 3190<br>(81.2%) | 1185<br>(85.0%) | 98 (74.2%) | 12022<br>(73.0%) |
| Never | 42 (0.4%) | 5 (0.1%) | 1 (0.1%) | 0 (0%) | 48 (0.3%) |
| Sometimes | 3416<br>(31.0%) | 729 (18.6%) | 208 (14.9%) | 34 (25.8%) | 4387<br>(26.6%) |
| **In the past 14 days have**<br>**you traveled to a US state**<br>**other than Washington**<br>**State?** |  |  |  |  |  |
| No | 9369<br>(85.0%) | 3631<br>(92.4%) | 1263<br>(90.6%) | 119 (90.2%) | 14382<br>(87.3%) |
| Yes | 1653<br>(15.0%) | 297 (7.6%) | 131 (9.4%) | 13 (9.8%) | 2094<br>(12.7%) |
| **In the past 14 days, have**<br>**you visited a country**<br>**other than the US?** |  |  |  |  |  |
| No | 10883<br>(98.7%) | 3910<br>(99.5%) | 1380<br>(99.0%) | 128 (97.0%) | 16301<br>(98.9%) |
| Yes | 139 (1.3%) | 18 (0.5%) | 14 (1.0%) | 4 (3.0%) | 175 (1.1%) |

\*2 were missing a response
†24 were missing a response
‡1008 were missing a response
§15 were missing a response
¶19 were missing a response

**Supplemental Table 6.**
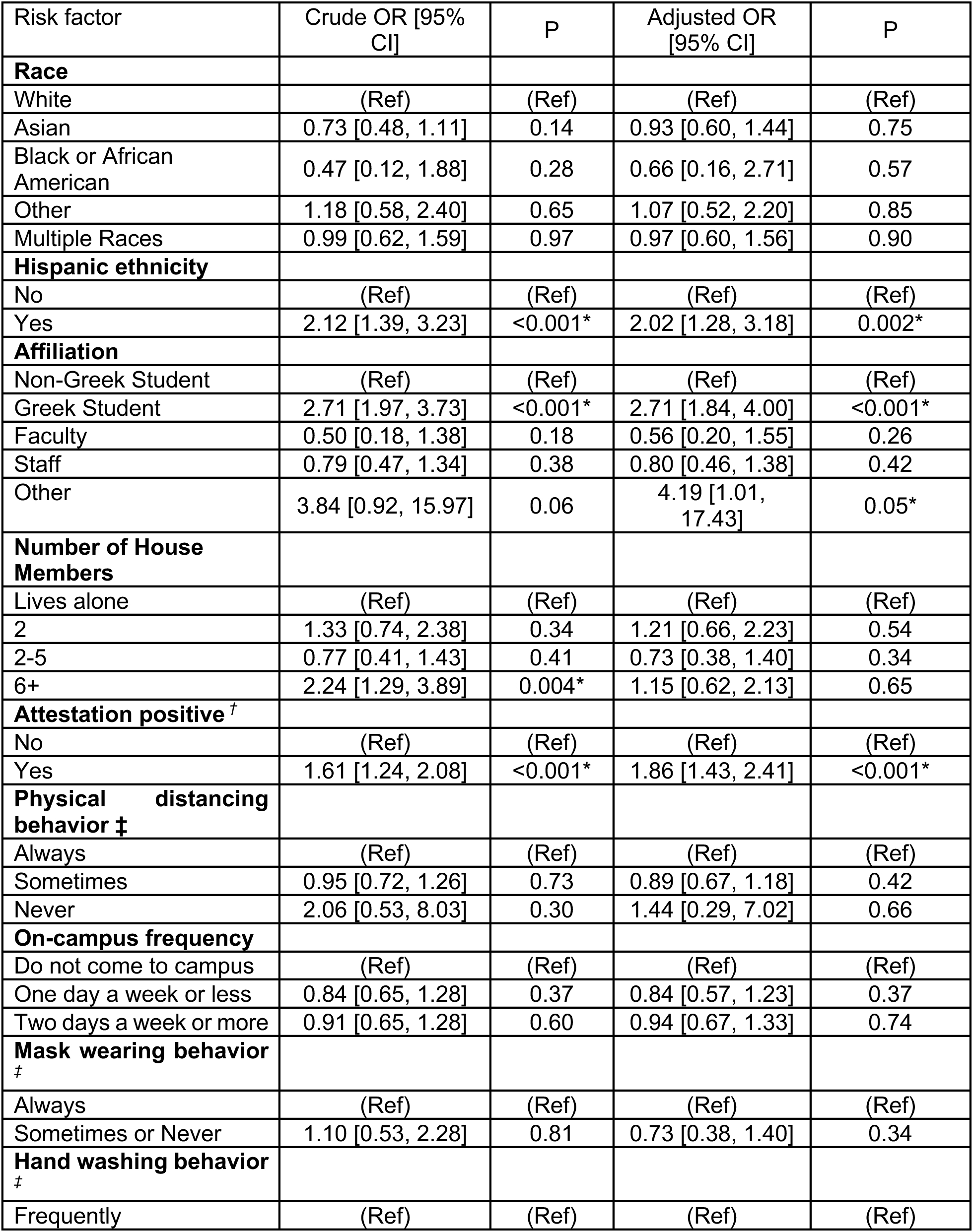
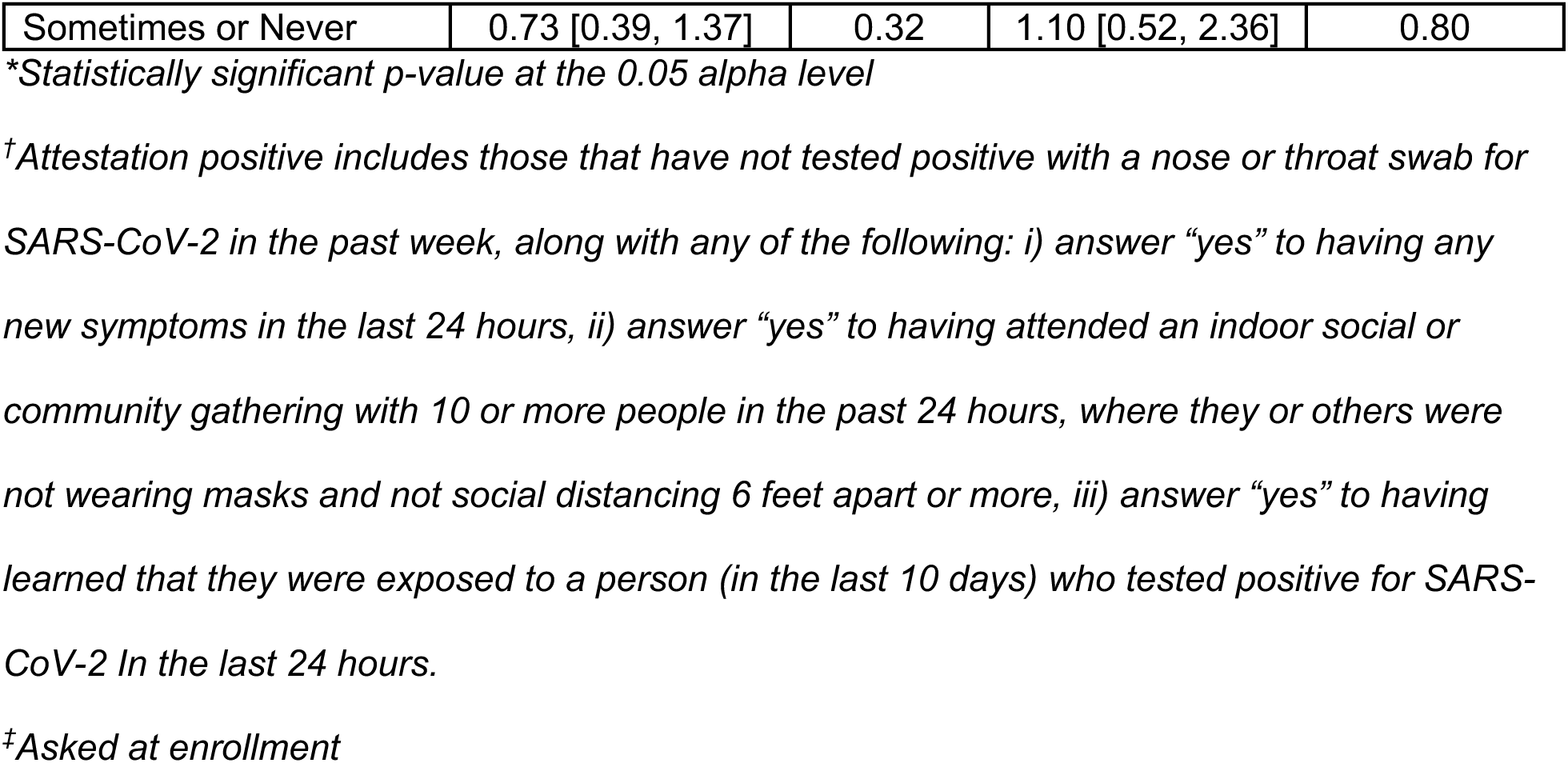
**Generalized Estimating Equation logistic regression model with robust variance**

| Risk factor | Crude OR [95% CI] | P | Adjusted OR [95% CI] | P |
| --- | --- | --- | --- | --- |
| **Race** |  |  |  |  |
| White | (Ref) | (Ref) | (Ref) | (Ref) |
| Asian | 0.73 [0.48, 1.11] | 0.14 | 0.93 [0.60, 1.44] | 0.75 |
| Black or African American | 0.47 [0.12, 1.88] | 0.28 | 0.66 [0.16, 2.71] | 0.57 |
| Other | 1.18 [0.58, 2.40] | 0.65 | 1.07 [0.52, 2.20] | 0.85 |
| Multiple Races | 0.99 [0.62, 1.59] | 0.97 | 0.97 [0.60, 1.56] | 0.90 |
| **Hispanic ethnicity** |  |  |  |  |
| No | (Ref) | (Ref) | (Ref) | (Ref) |
| Yes | 2.12 [1.39, 3.23] | <0.001* | 2.02 [1.28, 3.18] | 0.002* |
| **Affiliation** |  |  |  |  |
| Non-Greek Student | (Ref) | (Ref) | (Ref) | (Ref) |
| Greek Student | 2.71 [1.97, 3.73] | <0.001* | 2.71 [1.84, 4.00] | <0.001* |
| Faculty | 0.50 [0.18, 1.38] | 0.18 | 0.56 [0.20, 1.55] | 0.26 |
| Staff | 0.79 [0.47, 1.34] | 0.38 | 0.80 [0.46, 1.38] | 0.42 |
| Other | 3.84 [0.92, 15.97] | 0.06 | 4.19 [1.01, 17.43] | 0.05* |
| **Number of House Members** |  |  |  |  |
| Lives alone | (Ref) | (Ref) | (Ref) | (Ref) |
| 2 | 1.33 [0.74, 2.38] | 0.34 | 1.21 [0.66, 2.23] | 0.54 |
| 2-5 | 0.77 [0.41, 1.43] | 0.41 | 0.73 [0.38, 1.40] | 0.34 |
| 6+ | 2.24 [1.29, 3.89] | 0.004* | 1.15 [0.62, 2.13] | 0.65 |
| **Attestation positive <sup>†</sup>** |  |  |  |  |
| No | (Ref) | (Ref) | (Ref) | (Ref) |
| Yes | 1.61 [1.24, 2.08] | <0.001* | 1.86 [1.43, 2.41] | <0.001* |
| **Physical distancing behavior <sup>‡</sup>** |  |  |  |  |
| Always | (Ref) | (Ref) | (Ref) | (Ref) |
| Sometimes | 0.95 [0.72, 1.26] | 0.73 | 0.89 [0.67, 1.18] | 0.42 |
| Never | 2.06 [0.53, 8.03] | 0.30 | 1.44 [0.29, 7.02] | 0.66 |
| **On-campus frequency** |  |  |  |  |
| Do not come to campus | (Ref) | (Ref) | (Ref) | (Ref) |
| One day a week or less | 0.84 [0.65, 1.28] | 0.37 | 0.84 [0.57, 1.23] | 0.37 |
| Two days a week or more | 0.91 [0.65, 1.28] | 0.60 | 0.94 [0.67, 1.33] | 0.74 |
| **Mask wearing behavior <sup>‡</sup>** |  |  |  |  |
| Always | (Ref) | (Ref) | (Ref) | (Ref) |
| Sometimes or Never | 1.10 [0.53, 2.28] | 0.81 | 0.73 [0.38, 1.40] | 0.34 |
| **Hand washing behavior <sup>‡</sup>** |  |  |  |  |
| Frequently | (Ref) | (Ref) | (Ref) | (Ref) |
| Sometimes or Never | 0.73 [0.39, 1.37] | 0.32 | 1.10 [0.52, 2.36] | 0.80 |

*\*Statistically significant p-value at the 0.05 alpha level*
*†Attestation positive includes those that have not tested positive with a nose or throat swab for SARS-CoV-2 in the past week, along with any of the following: i) answer “yes” to having any new symptoms in the last 24 hours, ii) answer “yes” to having attended an indoor social or community gathering with 10 or more people in the past 24 hours, where they or others were not wearing masks and not social distancing 6 feet apart or more, iii) answer “yes” to having learned that they were exposed to a person (in the last 10 days) who tested positive for SARS-CoV-2 In the last 24 hours.*
*‡Asked at enrollment*

**Supplemental Table 7.** **Reported symptoms by SARS-CoV-2 test result**

|  |  | Positive<br>(n=265) | Negative<br>(n=29458) | Overall<br>(n=29783) | Percent<br>Positive |
| --- | --- | --- | --- | --- | --- |
| Reported<br>symptoms<br>within 7 days<br>prior to or at<br>time of<br>testing<br>(n=11116) | Muscle or<br>body aches | 54 (20.4%) | 2251 (7.6%) | 2313 (7.8%) | 2.3% |
|  | Difficulty<br>breathing | 18 (6.8%) | 1168 (4.0%) | 1191 (4.0%) | 1.5% |
|  | Chills | 36 (13.6%) | 1327 (4.5%) | 1365 (4.6%) | 2.6% |
|  | Cough | 74 (27.9%) | 3626 (12.3%) | 3711 (12.5%) | 2.0% |
|  | Fever | 52 (19.6%) | 1461 (5.0%) | 1518 (5.1%) | 3.4% |
|  | Loss of smell<br>or taste | 19 (7.2%) | 362 (1.2%) | 382 (1.3%) | 5.0% |
|  | Other | 134 (50.6%) | 8982 (30.5%) | 9147 (30.7%) | 1.5% |
|  | None | 49 (18.5%) | 13702<br>(46.5%) | 13771<br>(46.2%) | 0.40% |
|  | Missing | 55 (20.8%) | 4835 (16.4%) | 4896 (16.4%) | 1.1% |
| Reported<br>symptoms<br>within 7 days<br>after testing<br>(n=2897) | Muscle or<br>body aches | 42 (15.8%) | 611 (2.1%) | 657 (2.2%) | 6.4% |
|  | Difficulty<br>breathing | 19 (7.2%) | 344 (1.2%) | 364 (1.2%) | 5.2% |
|  | Chills | 20 (7.5%) | 375 (1.3%) | 397 (1.3%) | 5.0% |
|  | Cough | 26 (9.8%) | 794 (2.7%) | 825 (2.8%) | 3.2% |
|  | Fever | 35 (13.2%) | 414 (1.4%) | 453 (1.5%) | 7.7% |
|  | Loss of smell<br>or taste | 39 (14.7%) | 100 (0.3%) | 141 (0.5%) | 27.7% |
|  | Other | 51 (19.2%) | 1406 (4.8%) | 1462 (4.9%) | 3.5% |
|  | None | 83 (31.3%) | 18943<br>(64.3%) | 19065<br>(64.0%) | 0.44% |
|  | Missing | 99 (37.4%) | 7715 (26.2%) | 7821 (26.3%) | 1.3% |

**Supplemental Figure 3:**
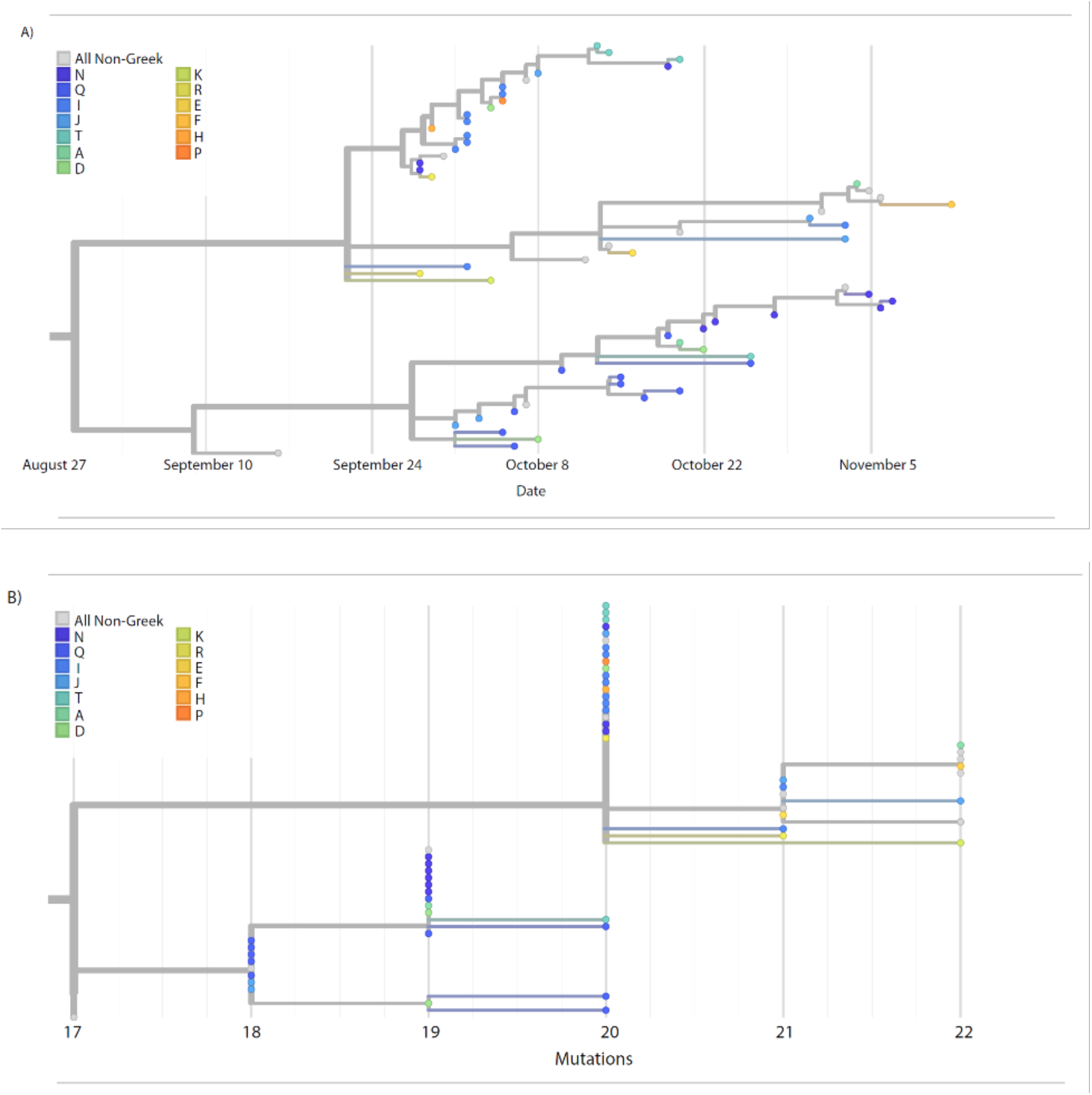
A) Detailed view of large cluster of SARS-CoV-2 samples collected by the study. Samples from Greek students are colored according to the house to which the students belong. Samples positioned along the x axis according to collection date. B) Identical to (A) with samples positioned along the x axis according to the number of mutations distinguishing them from the Wuhan/Hu-1 reference sequence.

**Supplemental Figure 4:**
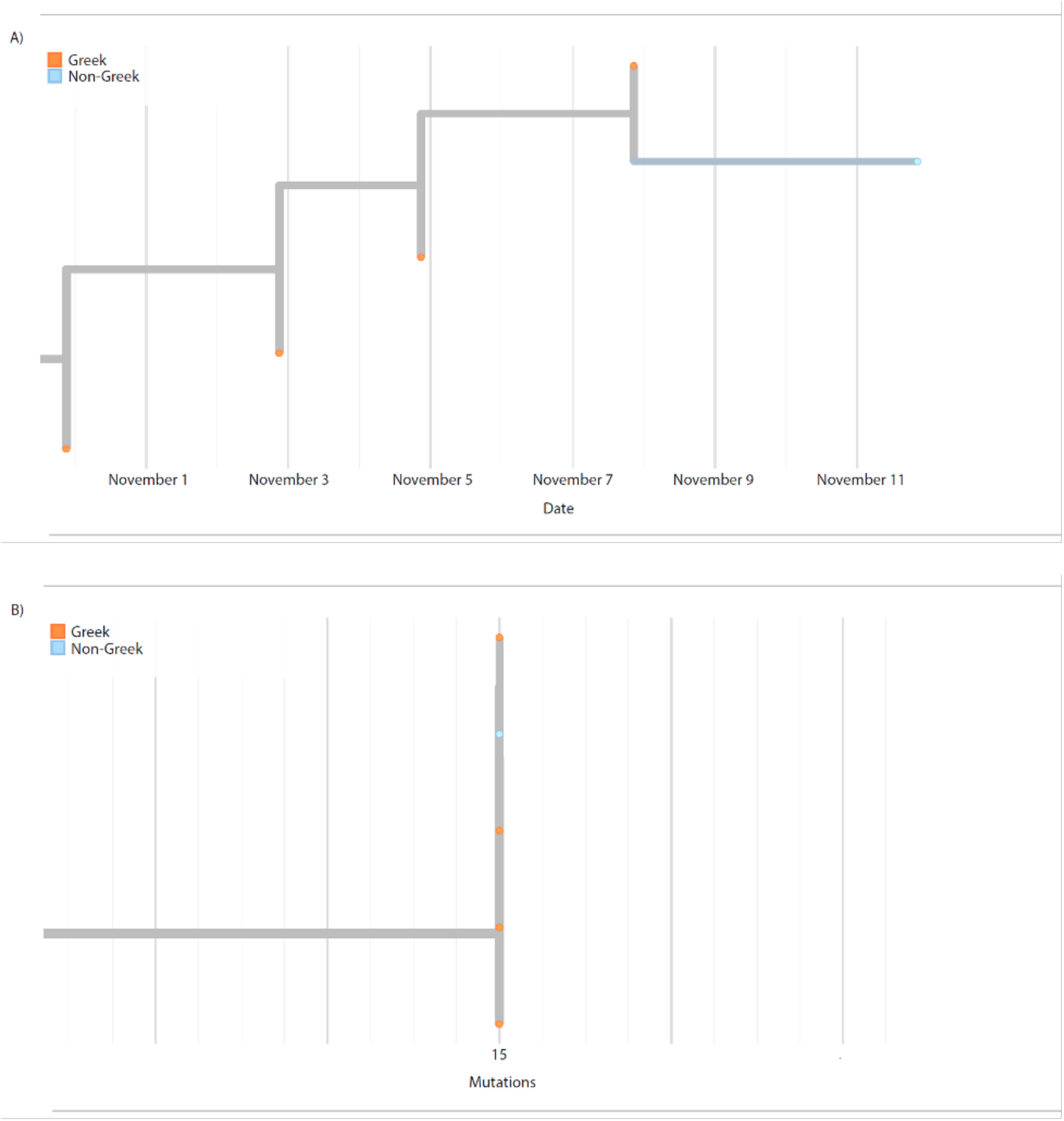
A) Detail of transmission cluster (boxed in *Figure 4A*) containing SARS-CoV-2 samples from 4 Greek students and one non-Greek student. Samples positioned along the x axis according to collection date. B) Identical to (A) with samples positioned along the x axis according to the number of mutations distinguishing them from the Wuhan/Hu-1 reference sequence.

**Supplemental Figure 5:**
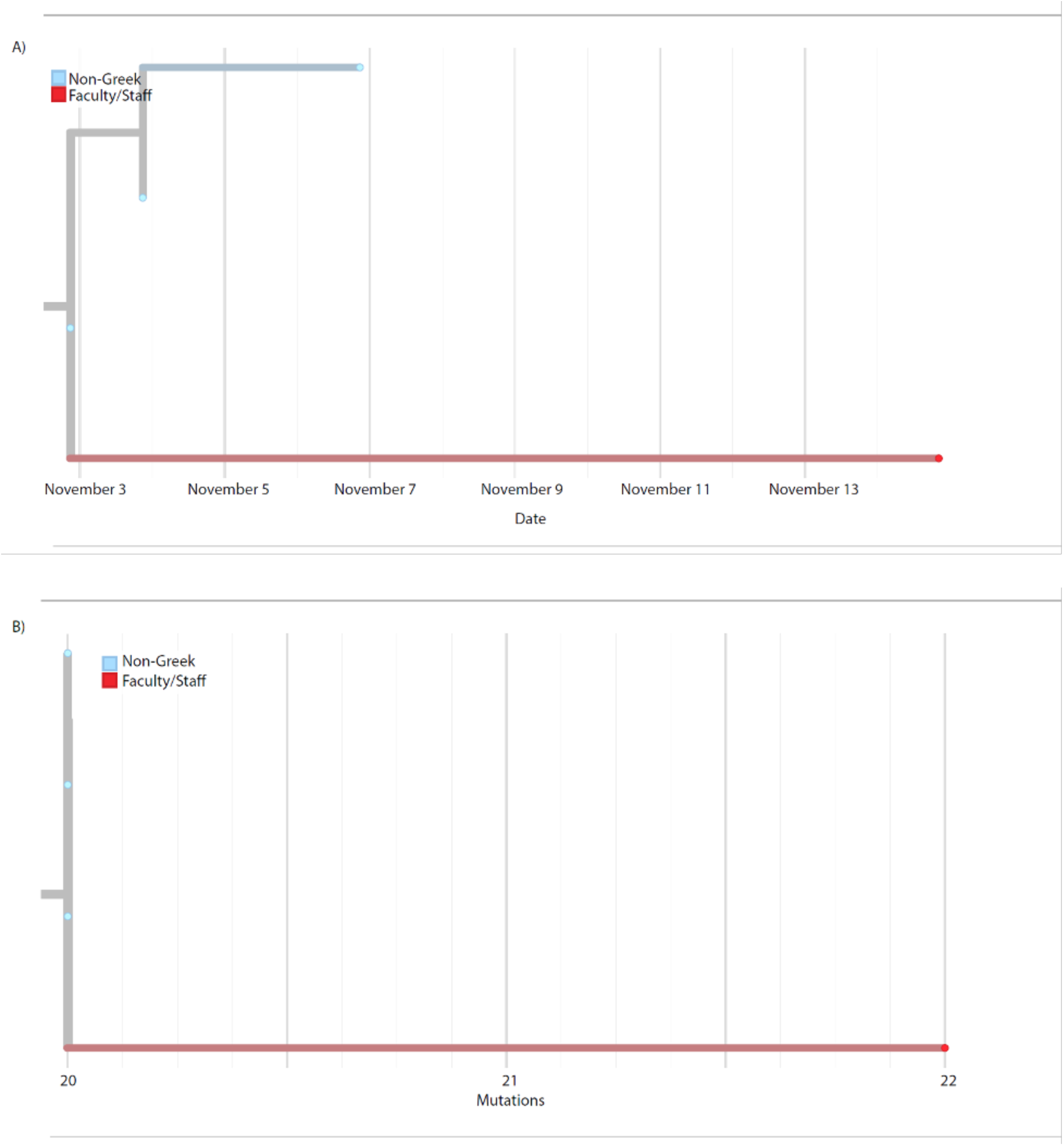
A) Detail of transmission cluster (boxed in *Figure 4A*) containing SARS-CoV-2 samples from 3 non-Greek students and one faculty/staff member. Samples are positioned along the x axis according to collection date. B) Identical to (A) with samples positioned along the x axis according to the number of mutations distinguishing them from the Wuhan/Hu-1 reference sequence.

**Supplemental Figure 6.**
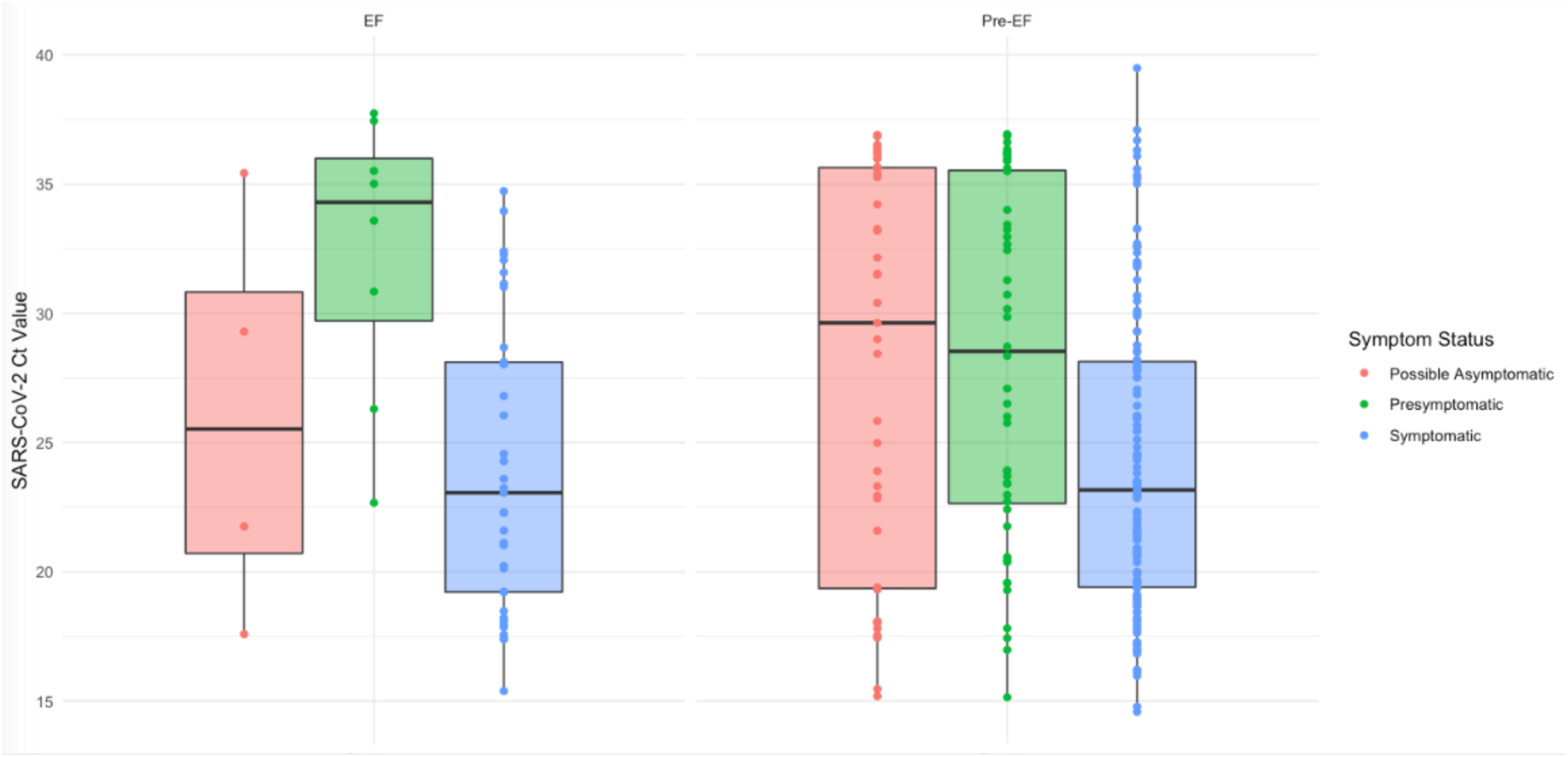
Ct values by symptom status: positive and inconclusive samples. Mid-study, we implemented an extraction-free testing method that yielded similar results, but Ct values from the two methods are not directly comparable. Cycle threshold (Ct) for samples (each represented by one dot) tested using our prior nucleic acid extraction testing method (‘Pre-EF’) and newer extraction-free laboratory method (‘EF’, implemented November 18, 2020) are shown here. Box plots show the median values and 25^th^ and 75^th^ percentiles, with vertical lines demonstrating the range of values.

## Appendices: Study Materials

1. Husky Coronavirus Testing Consent Form
2. Husky Coronavirus Testing Parental Consent Form
3. Eligibility Screening Questionnaire
4. Enrollment Questionnaire
5. Daily Attestation Survey
6. Study Protocol

## Supplemental Text

## Supplemental Methods

### Recruitment

Recruitment was conducted through university emails and newsletters, social media, flyers, announcements, physical signs, and links to the study website on a university webpage. For participants between 13-17 years of age, parent or guardian consent was required.

### Testing Procedures

#### In-person testing

Walk-up testing for individuals without a scheduled appointment was accepted, regardless of attestation or invitation status. Individuals enrolled onsite, either through smartphones or tablet. To minimize the possibility of contact with high-risk people during the testing process, individuals were tested in two physically separate groups. The first was for participants reporting symptoms or a high-risk exposure, and the second was for lower-risk study participants without these risk factors.

Swabs were collected by rotating the swab 360 degrees five times in each nostril and placing the swab in a dry tube with a barcode. The research staff scanned the barcode into the participant’s record. If participants tested positive/inconclusive and did not report symptoms within seven days prior to or following their test, they were sent a short follow-up survey about symptoms experienced after the positive test.

#### Mail-in swab kits

Mail-in swab kits were scheduled, delivered and picked up seven days per week [1][2]. For individuals receiving mail-in swab kits, the kits were delivered to the participant’s residence through a local courier service and picked up the same or next day. Swab kits contained a sealed anterior nasal swab, an instructional pamphlet, a dry tube with a barcode sticker, a biohazard bag, and a pre-addressed return polymailer bag [1–3]. Samples were returned in an International Air Transport Association-compliant Category B mailer within 48 hours of collection.

### Laboratory Methods

Swabs were transported dry, rehydrated and eluted in 1 mL of PBS or Tris-EDTA. 200 µL of eluate was extracted on the Magna Pure 96 using a DNA and Viral NA Small Volume Kit (Roche, 06543588001) with the universal small volume protocol, and eluted into 50 µL proprietary elution buffer. 200 µL of eluted anterior nares specimens collected after October 18, 2020 were extracted on the KingFisher Flex using the MagMAX Viral Pathogen II Nucleic Acid Isolation Kit with MagMAX™ Viral/Pathogen Ultra Enzyme Mix (Thermo Fisher A48383 and A42366) and eluted in 50 µL. After November 18, 2020 samples were prepared for PCR using an extraction-free method in which 50 µL of eluate in Tris-EDTA was heat treated for 30 min at 95°C used directly in RT-qPCR [3].

### Supplemental Results: COVID-19 Preventative Behavior

In the enrollment survey, participants reported frequency of COVID-19 prevention behaviors during the prior seven days (Supplemental Table 5). More than 93.6% and 97.4% reported frequent handwashing and mask wearing, respectively, while 52.0% reported frequently cleaning or disinfecting items and surfaces in their living or workspace, and 73.4% reported always social distancing (“In the past 7 days, how often did you try to stay 6 feet away from people who don’t live with you?”). Faculty/staff reported increased social distancing (81.4% and 85.2%) compared to students (69.3%). At enrollment, 245 (1.4%) of participants reported travel outside of the country in the prior 14 days, while 2421 (13.8%) reported traveling outside of the state during the same time frame.

